# Novel Genetic Locus Associated with Resistance to *M. tuberculosis* Infection: A Multi-Ancestry Genome-Wide Association Study

**DOI:** 10.64898/2026.03.06.26347614

**Authors:** Neel R. Gandhi, Matheus Fernandes Gyorfy, Mandar Paradkar, Nombuyiselo Jennet Mofokeng, Marina C. Figueiredo, Senbagavalli Prakash, Kamakshi Prudhula Devalraju, Qin Hui, Fay Willis, Vidya Mave, Bruno B. Andrade, Tumelo Moloantoa, Venkata Sanjeev Kumar Neela, Angela Campbell, Chang Liu, Alexandra Young, Marcelo Cordeiro-dos-Santos, Sanjay Gaikwad, Rajesh Karyakarte, Valeria C. Rolla, Afranio L. Kritski, Jeffrey M. Collins, N. Sarita Shah, James C.M. Brust, Vijaya Lakshmi Valluri, Sonali Sarkar, Timothy R. Sterling, Neil A. Martinson, Amita Gupta, Yan V. Sun

## Abstract

Understanding host susceptibility to *Mycobacterium tuberculosis* (*Mtb)* is critical for the development of new vaccines. Certain individuals “resist” becoming infected with *Mtb* despite intensive exposure; however, it is unknown whether there is a genetic basis for “resistance” to *Mtb* infection across populations. Here we conducted a genome-wide association study (GWAS) of resistance to *Mtb* infection by carefully characterizing exposure to TB patients among 4,058 close contacts in India, Brazil, and South Africa. 476 (12%) “resisters” remained free of *Mtb* infection despite substantial exposure to highly infectious TB patients. GWAS identified a novel chromosome 13 locus (rs1295104126) associated with resistance across the multi-ancestry meta-analysis. Comparing *Mtb*-infection to all uninfected contacts, irrespective of exposure, yielded a different locus on chromosome 6 (rs28752534), near the HLA-II region. These findings demonstrate a common genetic basis for resistance to *Mtb* infection across multi-ancestral cohorts with potential to elucidate novel mechanisms of protection from *Mtb* infection.

---

Tuberculosis (TB) is the leading infectious disease cause of death globally, with an estimated 10.7 million new diagnoses annually.^1^ The End TB Strategy set a goal of reducing global TB incidence to <10 cases per 100,000 population by 2035; however, this is not achievable with currently available TB control strategies.^2–4^ New tools to prevent new *Mycobacterium tuberculosis* (*Mtb*) infections in high incidence settings are needed.^2,3^ Yet, the development of biomedical interventions to prevent *Mtb* infection, such as a new vaccine, has been hampered by a poor understanding of host mechanisms that mediate susceptibility to *Mtb*.^5,6^ A greater understanding of the biologic mechanisms that allow specific individuals to “resist” *Mtb* infection would inform the development of new prevention modalities.

A genetic basis for “resistance to *Mtb* infection” has long been postulated.^7–10^ In numerous well-characterized events, individuals have been clearly exposed to *Mtb* by close contact with an infectious TB patient. These instances have included household or work settings (e.g., in naval ships or gold mines), living together before the index patient’s diagnosis.^11–15^ Across these studies, 5–20% of highly exposed contacts remained negative on tests of *Mtb* infection (e.g., interferon gamma release assays [IGRA] or tuberculin skin tests [TST]).^11–15^ It is unclear whether these individuals are truly not infected with *Mtb* or just do not produce an interferon gamma response; however, long-term follow-up data show that these individuals have a very low 10-year risk of developing TB disease or converting their IGRA/TST tests.^16^ These examples provide compelling evidence that host factors – not just the degree of exposure – influence whether an exposed individual becomes *Mtb*-infected or resists infection.

Difficulty in identifying cohorts of individuals resistant to *Mtb* infection have limited the rigorous study of resistance. These challenges include the inability to precisely diagnose *Mtb* infection, because IGRA and TST measure an immune response, not the *Mtb* bacteria. Similarly, *Mtb* exposure is often not carefully measured, raising the question of whether IGRA/TST-negative close contacts were sufficiently exposed. Differing criteria can influence the proportion of contacts identified as “resistant,”^15^ and vary the sensitivity or specificity of who is captured or misclassified as resistant.

Genome-wide association studies (GWAS) have revolutionized the study of genetic influences on rare and common diseases by enabling the large-scale identification of variants associated with a disease phenotype.^17^ Rather than being limited by *a priori* hypotheses, GWAS takes an “unbiased” approach facilitating the identification of loci not previously known to have a role in susceptibility.^18,19^ Over the past decade, GWAS has elucidated factors associated with susceptibility and pathogenesis in various infectious diseases, including hepatitis C virus, HIV, and malaria.^20–25^ Numerous GWAS have been carried out in TB over the past 15 years, but they have primarily focused on active TB disease.^26,27^

Early studies of resistance to *Mtb* infection utilized candidate gene and family linkage methodologies.^28–33^ Although some studies uncovered statistically significant associations, findings were limited by the genetic knowledge at the time or were not replicated.^34,35^ Further, these studies had limited resolution of genetic variants compared to GWAS.^36^ Two recent GWAS studies have investigated resistance to *Mtb* infection;^37,38^ however, a small sample size and a lack of measurement of *Mtb* exposure make it likely that specific variants and genes associated with resistance have not yet been identified.

We carried out a human GWAS study of resistance to *Mtb* infection by enrolling 4,058 close contacts of infectious TB index patients in India, Brazil, and South Africa. We carefully characterized exposure to the TB index patient and assessed the likelihood of the contact being resistant to *Mtb* infection. Based on this careful phenotyping, we used GWAS to identify genetic variants associated with resistance to *Mtb* infection.

## Results

The study enrolled 4,058 close contacts from India (n=1,658), Brazil (n=1,558), and South Africa (n=842), in collaboration with Regional Prospective Observational Research in TB (RePORT) consortia^39^ (**Table 1**, **Supplemental Table S1**). Among these participants, 2,385 (59%) were female and the median age was 27 years (IQR 15-43). Of those with exposure data available, the proportion who were highly exposed (slept in same bed/room, spent ≥5 hours indoors) to their index patient was 80% (2,644 of 3,310, **Table 1**). Sixty-five percent (2,261 of 3,453) of participants were exposed to highly infectious index patients (cavitary disease, AFB smear 2+/3+). Overall, 55% (1,896 of 3,428) of participants who had an IGRA test performed were positive and 54% (805 of 1,488) had a positive TST test (**Table 1** & **Supplemental Table S2**). The median time between the index patient’s diagnosis and last negative IGRA tested was 293 days (IQR 204-578 days).

**Table 1:** Baseline description of participants overall and by individual cohorts.

| Characteristics | Total<br>(N=4058) | Brazil Cohorts<br>(N=1558) | India Cohorts<br>(N=1658) | South Africa<br>Cohorts (N=842) |
| --- | --- | --- | --- | --- |
| Sex: Female | 2385 (59%) | 934 (60%) | 937 (57%) | 514 (61%) |
| Age: median years (IQR) | 27 (15-43) | 32 (16-47) | 26 (16-39) | 20 (12-39) |
| <5 | 98 (2.4%) | 66 (4.2%) | 27 (1.6%) | 5 (0.6%) |
| 5-12 | 637 (16%) | 212 (14%) | 209 (13%) | 216 (26%) |
| 13-20 | 816 (20%) | 222 (14%) | 378 (23%) | 216 (26%) |
| 21-30 | 719 (18%) | 228 (15%) | 349 (21%) | 142 (17%) |
| 31-40 | 666 (16%) | 274 (18%) | 333 (20%) | 59 (7.0%) |
| 41-50 | 498 (12%) | 236 (15%) | 217 (13%) | 45 (5.3%) |
| 51-60 | 348 (8.6%) | 187 (12%) | 104 (6.3%) | 57 (6.8%) |
| 61-70 | 191 (4.7%) | 100 (6.4%) | 32 (1.9%) | 59 (7.0%) |
| >70 | 85 (2.1%) | 33 (2.1%) | 9 (0.5%) | 43 (5.1%) |
| Exposure to Index patient |  |  |  |  |
| Slept in same bed | 554 (17%) | 182 (18%) | 237 (16%) | 135 (17%) |
| Slept in same room | 860 (26%) | 156 (15%) | 617 (42%) | 87 (11%) |
| Spent ≥5 hours indoors together | 1230 (37%) | 542 (52%) | 239 (16%) | 449 (55%) |
| None of the above | 666 (20%) | 154 (15%) | 367 (25%) | 145 (18%) |
| Not available | 748 | 524 | 198 | 26 |
| Index case infectiousness |  |  |  |  |
| High: Cavities present or AFB smear 2+/3+ | 2261 (65%) | 972 (62%) | 1136 (69%) | 153 (62%) |
| Medium: AFB smear scanty/1+, OR GeneXpert grade high or very high (no cavities) | 776 (22%) | 324 (21%) | 428 (26%) | 24 (9.7%) |
| Low: AFB smear negative OR GeneXpert grade low or very low (no cavities) | 416 (12%) | 262 (17%) | 86 (5.2%) | 68 (28%) |
| Not available | 605 | 0 | 8 | 597 |
| IGRA results |  |  |  |  |
| Positive | 1896 (55%) | 746 (48%) | 708 (65%) | 442 (56%) |
| Negative (>90 days) | 1311 (38%) | 794 (51%) | 220 (20%) | 297 (38%) |
| Negative (<90 days) | 221 (6.5%) | 16 (1.0%) | 160 (15%) | 45 (5.9%) |
| Not available | 630 | 2 | 570 | 58 |
| Days to last negative IGRA result: median (IQR) | 293 (204-578) | 241 (204-321) | 370 (88-874) | 847 (487-1063) |
| TST results |  |  |  |  |
| Positive | 805 (54%) | - | 805 (54%) | - |
| Negative (>90 days) | 154 (10%) | - | 154 (10%) | - |
| Negative (<90 days) | 529 (36%) | - | 529 (36%) | - |
| Not available | 2570 | 1558 | 170 | 842 |
| Days to last negative TST result: median (IQR) | 23 (7-89) | - | 23 (7-89) | - |

### Categorization of Resister and Infected Probability

Across all cohorts, 4000 participants had valid IGRA or TST test results (**Supplemental Figure S1**). Using IGRA/TST results, exposure, and index case infectiousness data, we created eight phenotypic categories to represent a spectrum from resistance to infection (**Figure 1**; see **Methods** and **Supplemental Methods** for full details). Participants with negative IGRA or TST results, high exposure (slept in same room or ≥5 hours spent indoors together) to a highly infectious index patient (cavitary disease or 2+/3+ smear grade) were considered to have the highest likelihood of being resistant to *Mtb* infection; they were placed in the Resister A, B, or C categories (**Figure 1**). The Uninfected A and B categories were comprised of participants with medium, low, or missing exposure or infectiousness data, and negative IGRA or TST results. Contacts with all positive IGRA or TST results, or a history of TB disease, were categorized as *Mtb*-infected (Infected A and B categories), whereas participants with discordant IGRA and TST results were classified as “Discordant” (**Figure 1**, **Table 2**).

**Figure 1.**
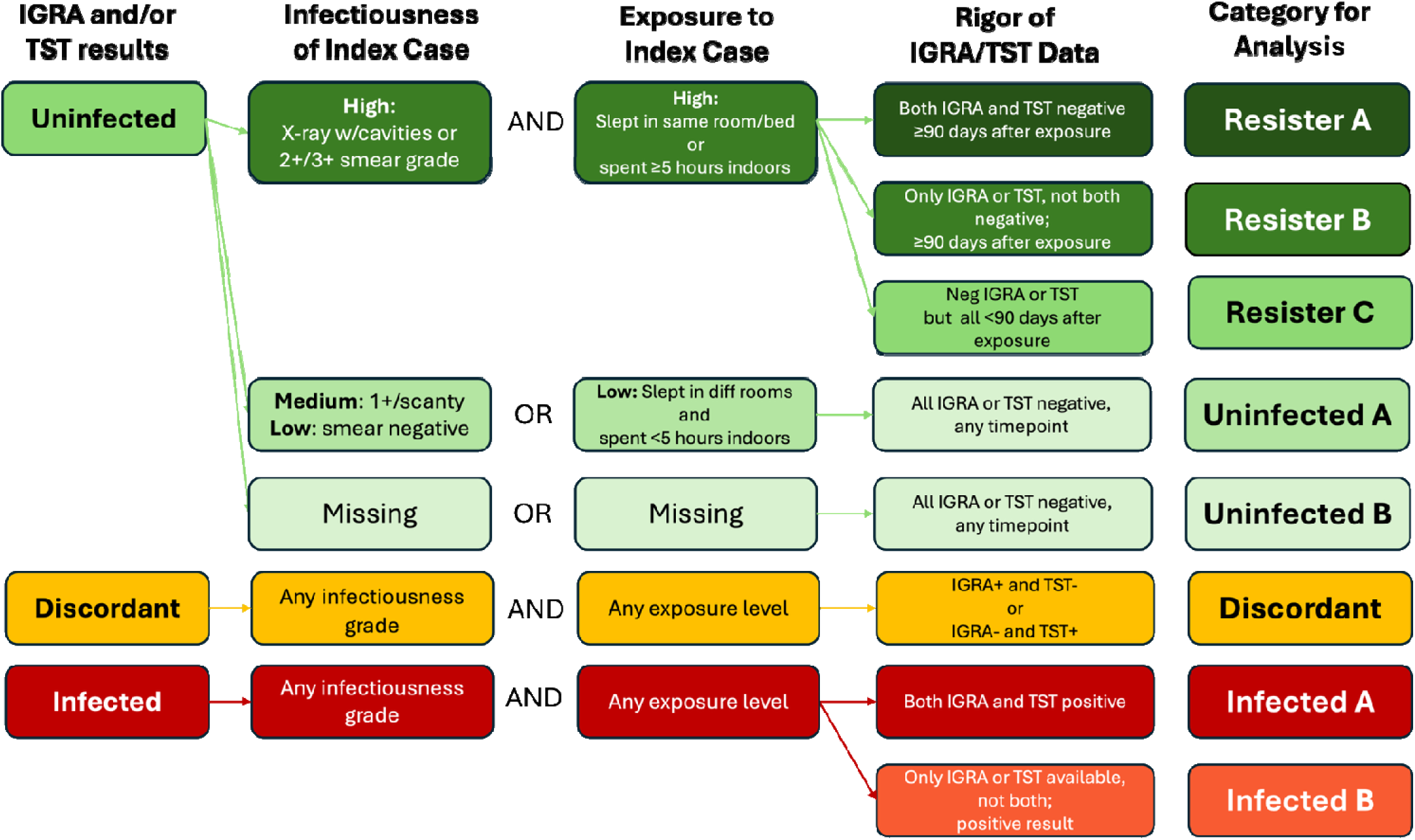
GWAS Resistance & Infection Hierarchy.

**Table 2:** Categories of likelihood of being resistant to *Mtb* infection and having *Mtb* infection, by country.

| Hierarchy | QFT/TST (Final) | Contact Exposure Level | Index infectiousness | Total<br>N=4000<br>N (%) | Brazil<br>N=1557<br>N (%) | India<br>N=1655<br>N (%) | South Africa<br>N=788<br>N (%) |
| --- | --- | --- | --- | --- | --- | --- | --- |
| Resister A | IGRA- & TST- (>90 days) | High | High | 35 (0.9%) | 0 ( 0%) | 35 (2.1%) | 0 ( 0%) |
| Resister B | IGRA- or TST- (>90 days) | High | High | 308 (7.7%) | 226 (15%) | 70 (4.2%) | 12 (1.5%) |
| Resister C | IGRA- or TST- (<90 days) | High | High | 133 (3.3%) | 5 (0.3%) | 122 (7.4%) | 6 (0.8%) |
| Uninfected A | IGRA- or TST- (any time) | Low | Medium/Low | 672 (17%) | 337 (22%) | 279 (17%) | 56 (7.1%) |
| Uninfected B | IGRA- or TST- (any time) | Unknown or Missing | Unknown or Missing | 504 (13%) | 222 (14%) | 36 (2.2%) | 246 (31%) |
| Discordant | IGRA+ & TST- or<br>IGRA- & TST+ | Any/Unk | Any/Unk | 357 (8.9%) | 7 (0.4%) | 329 (20%) | 21 (2.6%) |
| Infected B | IGRA+ OR TST+ | Any/Unk | Any/Unk | 1436 (36%) | 724 (46%) | 384 (23%) | 328 (42%) |
| Infected A | IGRA+ & TST+ | Any/Unk | Any/Unk | 555 (14%) | 36 (2.3%) | 400 (24%) | 119 (15%) |

Using these phenotypic categories, we found 476 (12%) participants were highly likely to be resistant (Resister A/B/C), consistent with previous studies of resistance to *Mtb* infection.^14,15,40^ An additional 1,176 (29%) participants were IGRA or TST negative, but it was unclear if they were resistant due to insufficient exposure (Uninfected A-B; **Table 2 Supplemental Table S3**).

The resisters were predominantly from India (n=227) and Brazil (n=231). The number of resisters from South Africa (n=18, 2.3%) is likely underestimated because index patients were diagnosed using the Xpert MTB/RIF assay and data to infer infectiousness (chest x-ray, AFB smear grade) were not collected (**Table 2**, **Supplemental Table S4)**; nonetheless, the ancestral diversity across the Indian and Brazilian cohorts provided a robust opportunity to still identify genetic variants in a multi-ancestry and admixed population in the GWAS for resistance.

Differences between the resister and infected categories based on age, sex, exposure, and infectiousness are presented in **Supplemental Table S5**.

### Genetic and Principal Components Analysis

Among the 4,058 participants enrolled, the GWAS assay was performed for 3,959 (98%; **Supplemental Figure S1**). After GWAS data quality control, 3,889 participants were included in the final genetic analyses (1,631 India, 1,527 from Brazil, 731 South Africa). Principal Components Analysis (PCA) demonstrated strong evidence of a multi-ancestry and admixed population, including African, European, Asian, and indigenous South American ancestries (**Supplemental Figure S2**).

### Genetic Markers Associated with Resistance to *Mtb* Infection

The GWAS for resistance compared participants with a high likelihood of being resistant (Resisters A/B/C) to all others (Uninfected A/B, Discordant, Infected A/B). Among Indian participants, 6,172,107 variants with a minor allele frequency (MAF) of ≥5% were used in the GWAS. We found one statistically significant association with a locus on chromosome 13 (SNP rs11843280, OR=2.53, 95%CI 1.83–3.50, p=1.71×10^-8^; **Supplemental Figure S3a**). (The odds ratios [OR] represent the difference in risk of having the outcome of interest [e.g., resistance] between the effect allele and reference allele at a given SNP [alleles presented in **Supplemental Table S6a].)**

For the Brazil analysis, we examined 6,931,279 SNPs with MAF of ≥5%. No loci achieved the genome-wide significance threshold of p<5×10^-8^ (**Supplemental Figure S3b**). However, the significant locus from the India GWAS had the same direction of effect in the Brazil cohort (SNP rs11843280, OR=1.74, 95%CI 1.26–2.40, p=0.0007, **Supplemental Table S6a**, **Supplemental Figure S4a**).

A meta-analysis combining the India and Brazil cohorts yielded an inflation factor of 1.0 (**Supplemental Figure S5a**). We identified the same locus on chromosome 13, with multiple statistically significant SNPs (highest association: SNP rs1295104126, OR = 0.47, 95% CI 0.37 – 0.59, p = 6.73×10^-11^; **Figure 2, Supplemental Table 5a**). The associated region on chromosome 13 exhibited high linkage disequilibrium (LD). There was a high correlation of genetic effects within the same region, highlighting that the SNPs identified were markers for the causal effect of the locus region (r^2^=0.89 between SNPs rs1295104126 and rs11843280, the top variants in meta-analysis and Indian analysis).

**Figure 2:**
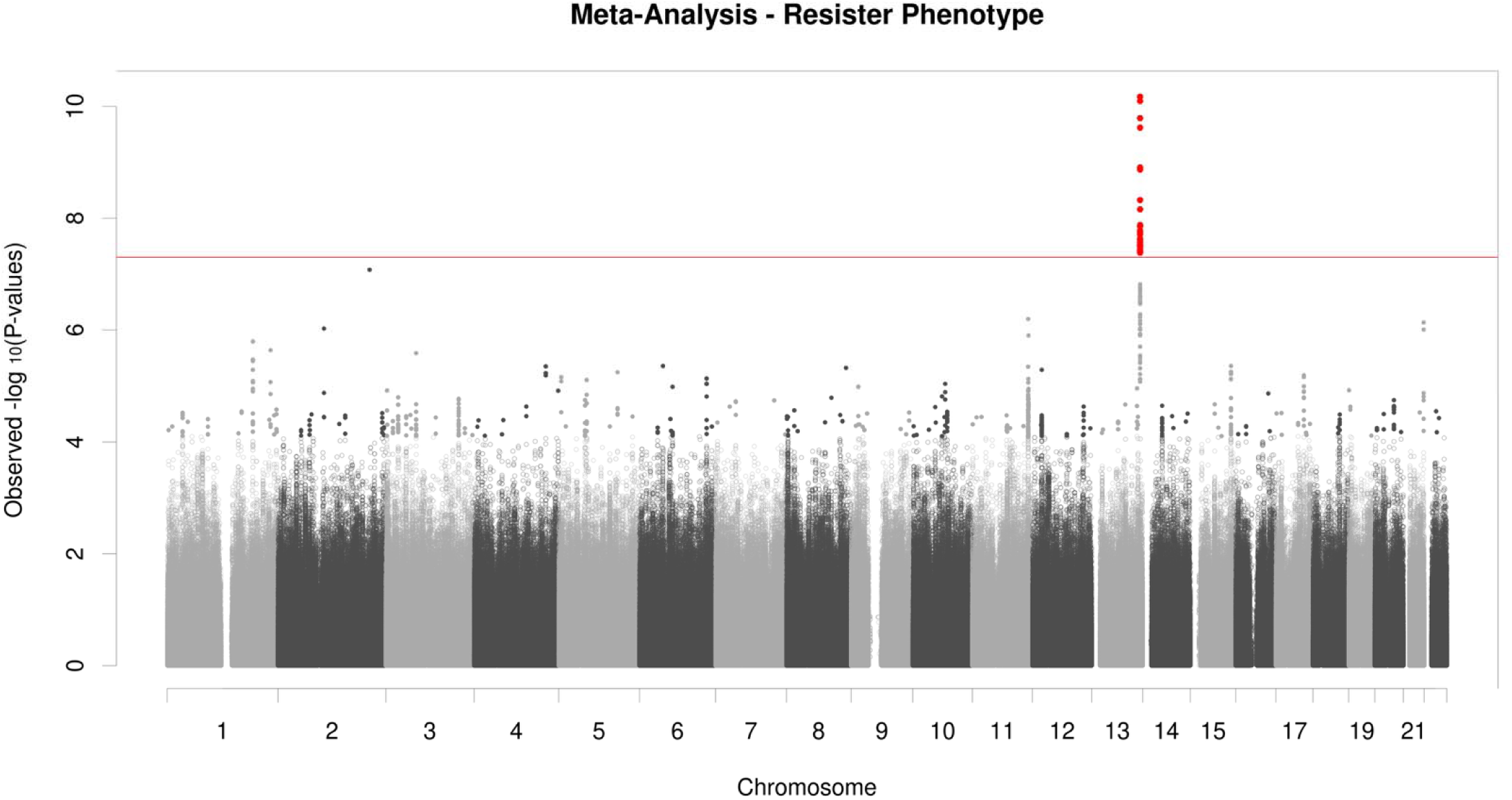
Genetic markers associated with Resistance to *Mtb* infection; Meta-analysis of GWAS from India and Brazil cohorts: One statistically significant locus on chromosome 13: rs1295104126 (OR = 0.47, 95% CI 0.37 – 0.59, p = 6.73×10^-11^)

Among the sensitivity analyses performed, Sensitivity Analysis #1 varied the control group to only Infected A-B and Discordant (**Supplemental Table S7a**); it showed that SNP rs1295104126 remained significant with a similar effect size but lower precision (OR=0.47, 95%CI 0.37–0.60, p=5.22×10^-10^; **Supplemental Table S7a, Supplemental Figure S6a**). In Sensitivity Analysis #2, the resister group (“cases”) was varied to include those who were missing exposure or infectiousness data (i.e., Uninfected B added to Resister A/B/C, **Supplemental Table S7a**). We found the effect size diminished and p-value was no longer genome-wide significant (OR=0.66, 95%CI 0.55–0.78, p=1.69×10^-6^). (See **Supplemental Results** for full discussion of sensitivity analyses results.)

The statistically significant locus on chromosome 13 was located in an intergenic region of the *MYO16* gene, which encodes for an unconventional myosin protein and is postulated to function as a regulatory subunit for serine/threonine phosphatase-1.

### Genetic Markers Associated with *Mtb* Infection

The GWAS for *Mtb* infection analysis included participants from India, Brazil, and South Africa. The Indian analysis used 6,172,107 SNPs, the Brazilian analysis used 6,931,279 SNPs, and the South African analysis used 8,478,562 SNPs. No individual country analysis had statistically significant associations at the genome-wide p<5×10^-8^ threshold.

The meta-analysis combining participants from all three countries, however, had a statistically significant locus on chromosome 6 (highest association SNP rs28752534, OR=0.69, 95%CI 0.62–0.78, p=2.06×10^-10^, inflation factor 1.001; **Figure 3, Supplemental Figure S5b**). (OR represents the difference in risk of having *Mtb* infection between the effect allele and reference allele at the SNP [alleles in **Supplemental Table S6b**].)

**Figure 3:**
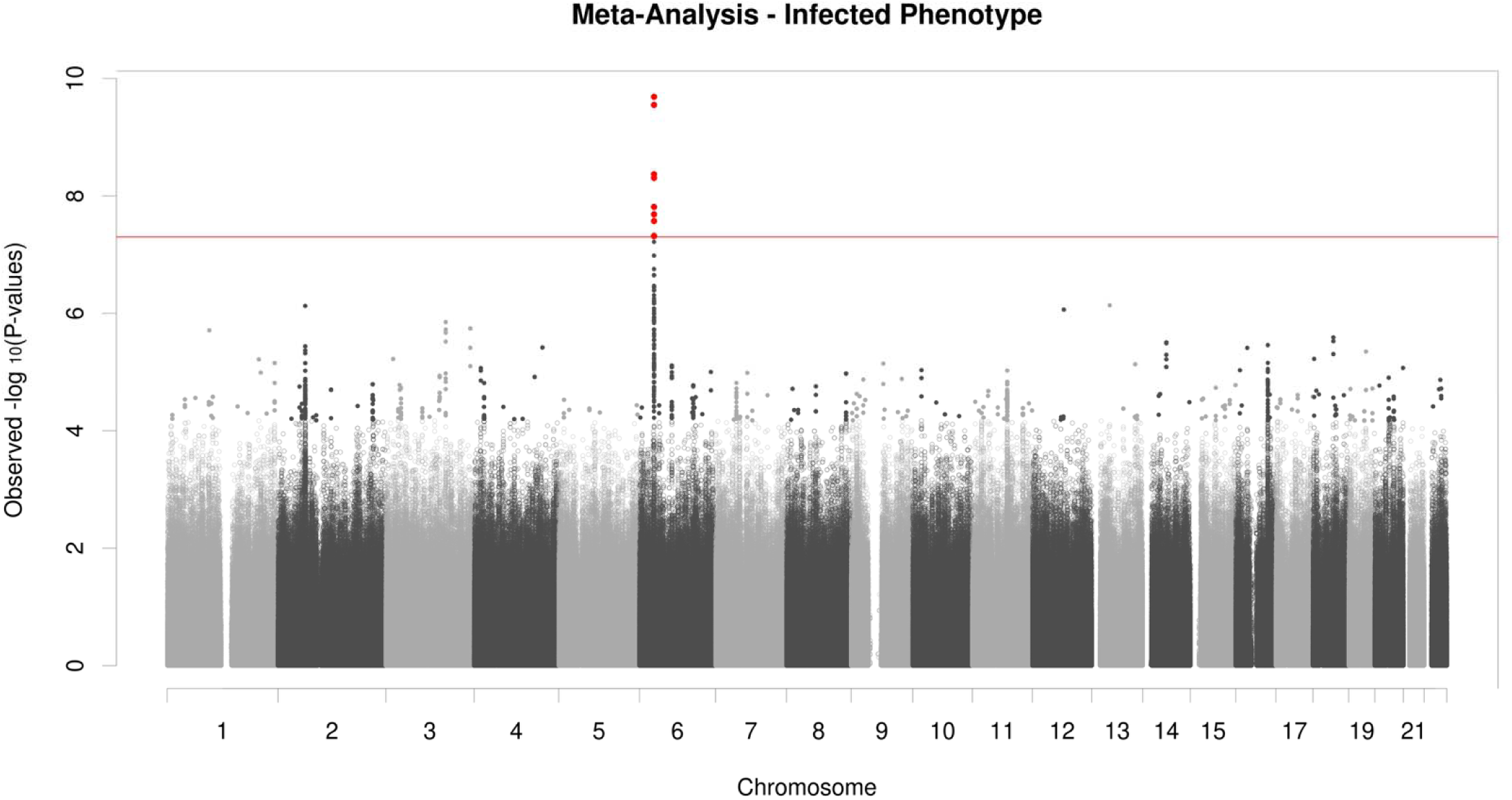
Genetic markers associated with *Mtb* infection; Meta-analysis of GWAS from India, Brazil, and South Africa cohorts: One statistically significant locus on chromosome 6 at rs28752534 (OR = 0.69, 95% CI 0.62 – 0.78, p = 2.06×10^-10^)

Sensitivity analysis #3 added participants from the Discordant IGRA/TST category to the Infected “case” group (i.e., Infected A-B plus Discordant; **Supplemental Table S7b**). This sensitivity analysis showed the same chromosome 6 locus was statistically significantly associated, but with a lower effect size (SNP rs28752534, OR=0.72, 95%CI 0.64–0.81, p=3.88×10^-8^; **Supplemental Figure S6b**). (See **Supplemental Results** for full discussion of sensitivity analysis results.)

This statistically significant locus is in a well-known intergenic region between genes HLA-DRB1 and HLA-DQB1, replicating findings of previous genetic studies of *Mtb* infection.

## Discussion

In this study, we sought to identify genetic predictors of resistance to *Mtb* infection. We rigorously characterized close contacts from diverse ancestries in India, Brazil, and South Africa to identify individuals with high exposure to *Mtb* who remained uninfected. We found a new chromosome 13 locus associated with resistance across multiple ancestral backgrounds in our GWAS for resistance, comparing uninfected individuals with high exposure to *Mtb* infection with all others. Interestingly, when we conducted the GWAS for *Mtb* infection, comparing *Mtb*-infected participants to all others, we identified a canonical HLA-II locus on chromosome 6 that has previously been associated with *Mtb* infection. These findings suggest that rigorous characterization of resister status based on exposure to infectious TB reveals a different biological mechanism of protection than genetic studies that compare *Mtb*-infected individuals to all uninfected persons without consideration of degree of exposure. Future research elucidating the underlying biological pathways associated with the novel chromosome 13 marker may reveal new mechanisms of protection against *Mtb* infection.

TB is characterized by a spectrum of infection and disease states following exposure. One recognized challenge in GWAS studies of TB has been clearly characterizing a discrete clinical phenotype for which genetic predictors are sought.^41^ This is true in studies of resistance to *Mtb* infection as well. We previously demonstrated that the individuals categorized as resisters vary based on the stringency used for exposure and IGRA/TST testing.^15^ In this study, we sought to minimize misclassification in the resister group by requiring they meet all of the following criteria: a very high index patient infectiousness, high degree of exposure to the infectious patient, and rigorous assessment of *Mtb* infection evaluating all IGRAs or TSTs performed. Despite the high threshold used, we still identified 476 (12%) resisters; this proportion is consistent with the existing resistance to *Mtb* infection literature.^14,15,40,42,43^ Further, our GWAS findings demonstrated that the genetic loci identified differ based on whether we utilize a rigorous resister definition or whether we simply use a definition of *Mtb* infection versus those not infected, without taking infectiousness and exposure into account.

Our GWAS for resistance analysis uncovered SNPs in a statistically significant locus close to the *MYO16* gene (rs1295104126) on chromosome 13. This locus was statistically significant among the Indian participants and remained significant when Brazilian participants, who had considerable genetic ancestral diversity, were added in meta-analysis. Our sensitivity analyses suggest that this locus is specifically associated with the stringent, highly exposed resister phenotype we characterized for this study. The effect size and strength of association both diminish when less exposed, uninfected groups were included in sensitivity analysis, suggesting greater misclassification of non-resisters into the resister group and dilution of the signal of association with resistance. In fact, as seen in our GWAS for *Mtb* infection, inclusion of all uninfected participants without careful efforts to minimize misclassification of non-resisters led to an entirely different locus of association.

The identified locus on chromosome 13 is a newly reported region in the TB-focused genetic literature; as such, it should be replicated and further investigated to better understand what molecular functions may relate to resistance to *Mtb* infection. The *MYO16* gene encodes myosin XVI, an unconventional member of the myosin family of actin-based motor proteins involved in various intracellular processes.^44,45^ Myosin XVI is found predominantly in the nervous system, but is also present in peripheral tissues, including the lung. Through its interaction with the WAVE Regulatory Complex it directs the remodeling of the cell’s internal skeleton.^46^ This role in cytoskeletal rearrangement could have implications in processes like phagocytosis and vesicle trafficking.^46,47^ Polymorphisms in the *MYO16* gene have previously been associated with neuropsychiatric diseases, such as schizophrenia or autism.^48,49^ Studies have also linked *MYO16* with illnesses in other organ systems, such as lupus and autoimmune diseases.^50^ The evidence for the presence of an intracellular role for *MYO16* highlights the need for additional studies to identify its role in host response to *Mtb*.

Our GWAS for *Mtb* infection analysis found a statistically significant locus in a well-known intergenic region between genes HLA-DRB1 and HLA-DQB1 (SNP rs28752534). Loci from this region have been associated with active TB disease and *Mtb* infection previously.^27,51–53^ An RNA-seq analysis demonstrated strong associations between HLA-DQA1 and HLA-DRB1 and the combined TB disease/*Mtb* infection phenotype.^54^ A systematic review and meta-analysis, including cohorts of European, Asian, and African ancestries, found that HLA class II genes, like HLA-DQB1, have demonstrated statistically significant protection against pulmonary TB disease.^55^ The fact that our GWAS for *Mtb* infection replicated the previously described associations is reassuring for comparability between our study population and others globally.

A strength of our study was the ancestral diversity represented by assembling cohorts across Asia, Africa, and South America, as demonstrated by the principal component analysis (**Supplemental Figures S2a-2c**). The Indian and South African participants represented South Asian and African populations well. The Brazilian participants represented admixture of African, European, Asian, and indigenous South American ancestries, consistent with Brazil’s cultural history. We performed each GWAS analysis by country to identify any potentially associated loci. Our meta-analyses across countries then determined whether our findings are robust across multiple ancestries. The global burden of TB continues to disproportionately impact countries across these continents, underscoring the potential generalizability and impact of our study findings.

Our study was subject to several limitations. First, there is no consensus definition for resistance nor minimum exposure needed to establish *Mtb* infection, limiting cross-study comparability. However, we have provided a systematic and transparent methodology to allow others to replicate a similar approach (**Figure 1**, **Table 2**). Further, our sensitivity analyses using alternate definitions provide support for our findings. Second, it is unclear whether persistently TST/IGRA-negative individuals are truly uninfected or have non-interferon-gamma t-cell responses to *Mtb*. However, participants in a Ugandan study who were persistently TST-negative over the first 6-12 months after exposure to an index case, were followed for an average of 9.5 years;^16^ nearly 85% of participants remained TST and IGRA negative and free of TB disease. These data suggest that persistent TST-negativity is durable in the vast majority of individuals and associated with low risk of TB disease, despite living in a high TB incidence setting. Third, not all data elements were available in every cohort included in our study. The major consequence of this was that missing index patient infectiousness data resulted in low numbers of Resisters A/B/C participants from our South African cohorts, preventing us from including them in the GWAS for resistance analysis (**Table 2**). Nonetheless, the genetic diversity seen across the Brazilian and Indian cohorts (**Supplemental Figure S2)**, provides support that our GWAS for resistance findings are robust across multiple cohorts, geographic regions, and ancestries.

A genetic basis to resistance to *Mtb* infection has long been postulated. By assembling a large multi-ancestry and admixed cohort and carefully characterizing their exposure to infectious TB, our study was able to identify a population of individuals with a high likelihood of resistance to *Mtb* infection and use GWAS analysis to identify a novel genetic locus associated with resistance. Future studies examining potential biologic mechanisms associated with this locus hold promise for elucidating new mechanisms of protection against *Mtb* infection.

## Methods

### Study Population

We conducted an observational study among close contacts of pulmonary TB patients enrolled in nine cohorts from 10 research centers across India, Brazil, and South Africa, in collaboration with RePORT consortia (**Supplemental Table S1**).^39^ Participants of all ages were eligible if exposed to a microbiologically confirmed index patient (e.g., positive culture, Xpert). The minimum exposure for eligibility was living in the same house for ≥1 month or, for non-household close contacts, spending ≥4 hours together per week after the index patient developed TB symptoms. Close contacts living with HIV were excluded since the impact of HIV on immune responses may lead to false-negative IGRA/TST results. (Close contacts of HIV co-infected TB index patients were still eligible, as long as the contact was HIV-negative.)

### Clinical Data and Characterization of Resister Phenotype

We created phenotypic categories characterizing the likelihood of participants being resistant to *Mtb* infection (“resister”) or being *Mtb*-infected. We harmonized data related to index patient infectiousness (e.g., chest x-ray, smear microscopy), contact exposure (e.g., hours spent indoors, sleeping arrangements), and the results and timing of all IGRA and/or TST tests performed from each cohort. (Because many cohorts began enrollment before this study, data availability varied across cohorts, **Supplemental Table S4**).

We created eight phenotypic categories representing a hierarchy of resistance and infection (**Figure 1**, **Supplemental Methods** for full details). Participants with negative results for all IGRA or TST tests were categorized into five categories reflecting differing probabilities of being resistant to *Mtb* infection. Participants with high exposure (slept in the same room or ≥5 hours spent indoors together) to a highly infectious index patient (cavitary disease or 2+/3+ smear grade) were considered to have the highest likelihood of being resistant to *Mtb* infection. These participants were placed in the Resister A, B, and C categories (**Figure 1**). The timing of IGRA/TST results was also assessed to account for potential IGRA/TST conversion; the Resister A-B categories included participants with negative IGRA/TST results >90 days after exposure. The Uninfected A and B categories were comprised of participants with medium, low, or missing exposure or infectiousness data, in whom it was unclear whether they had sufficient *Mtb* exposure to cause infection. Contacts with positive results for all IGRA or TST tests performed, or a history of TB disease, were categorized as *Mtb*-infected (Infected A-B categories). Participants with discordant IGRA and TST results were classified as “Discordant” (**Figure 1**, **Table 2**).

We collapsed these discrete categories into four main groups for the principal analyses: High likelihood of resistance to *Mtb* infection (Resisters A/B/C); Not infected but unclear if resistant (Uninfected A-B); Discordant; and *Mtb*-infected (Infected A-B, **Supplemental Table S3**). The detailed hierarchy, however, also allowed for different groupings in sensitivity analyses to account for uncertainties in determining *Mtb* infection status and degree of exposure (**Supplemental Table S7**).

### GWAS Design, Genotyping, Quality Control, and Imputation

We conducted GWAS analyses using two analytic approaches to identify genetic markers of resistance. The GWAS for resistance compared those with a high likelihood of being resistant (Resisters A/B/C) to all others (Infected A-B, Discordant, Uninfected A-B). The GWAS for *Mtb* infection compared those with certainty of having *Mtb* infection (Infected A-B) to all others (Discordant, Resisters A/B/C, Uninfected A-B; **Supplemental Tables S7a & S7b**).

We also conducted sensitivity analyses. For the GWAS for resistance, Sensitivity Analysis #1 varied the control group (i.e., only Infected A-B/Discordant; **Supplemental Table S7a**). Sensitivity Analysis #2 varied the resister group (“cases”) to account for those who were missing exposure or infectiousness data (i.e., Uninfected B added to Resister A/B/C). For the GWAS for *Mtb* infection, Sensitivity Analysis #3 examined the influence of including the Discordant IGRA/TST category into the Infected group (i.e., Infected A-B plus Discordant; **Supplemental Table S7b**).

Genotyping was performed using the Illumina Global Screening Array v3 (Illumina, San Diego, USA). Positive control and duplicated samples were included to ensure reproducibility. Sample genotype data were examined for adequate call rate (passing threshold >95%). Genotyping data were compared to demographic and phenotypic categorization data for quality control procedures; samples with a mismatch between genotype-derived gender and demographic gender, and duplicated samples were removed.

Genotype data were uploaded to Imputation Server (<u>imputation.biodatacatalyst.nhlbi.nih.gov</u>) for imputation with TOPMed Reference Panel using Minimac4. The imputed variants, in GRCh38, were then filtered by imputation quality R^2^ > 0.3 and used in GWAS analysis. We focused on common variants (MAF ≥ 5%) given our overall sample size and power considerations, and we also excluded variants with Hardy-Weinberg Equilibrium (HWE) p-value below 1×10^-4^.

### Statistical, Graphical, and Annotation Methods

Statistical analysis was performed using a generalized linear mixed model (GLMM) with penalized quasi-likelihood (PQL) approximation in the GENESIS software. We acquired Principal Components (PCs) through GENESIS PC-AiR function. The outcome variable for the GWAS for resistance analyses was <u>resistance</u> to *Mtb* infection (Resisters A/B/C) and for the GWAS for *Mtb* infection analyses was *Mtb* infection (Infected A-B). The predictor variable was the genotype of each SNP. The covariates included were age, sex, and the top 10 PCs to account for potential confounding factors, sample relatedness, and ancestry. Study samples’ genotype data were merged with the 1000 Genome Project reference panel to visualize ancestries,^56^ and non-autosomal chromosomes were not included.

GWAS analyses were carried out for participants from each country individually; data from across countries were later combined in a meta-analysis. A genome-wide significance threshold of p-value <5×10^-8^ was used to identify significant associations, and the locus was defined by a window of 250,000 base-pairs on each side of a sentinel SNP. Odds ratios (OR) represented the difference in risk of having the outcome of interest (e.g., resistance in GWAS for resister, infection in GWAS for *Mtb* infection) between the effect allele and reference allele at each SNP; alleles for SNPs with greatest statistical significance for each analysis are presented in **Supplemental Table S6**. An OR >1.0 indicated that individuals with the effect allele have a higher likelihood of the outcome of interest (e.g., resistance, *Mtb* infection); OR <1.0 indicates that individuals with the effect allele have a lower likelihood of the outcome of interest. The genomic inflation factor (l) was calculated by dividing the median of the observed χ^2^ test statistic by the expected median of the corresponding χ^2^ distribution. Post-analysis annotations were made using the MAGMA tool in FUMA’s SNP2GENE function (v1.5.2).

## Ethical Considerations

The study was approved by the institutional review board at Emory and all collaborating institutions (full details in **Supplemental Methods**).

## Acknowledgements and Funding sources

The study was funded by a grant from the US National Institutes of Health-National Institute of Allergy and Infectious Disease (NIH/NIAID): R01AI139406 (MPIs: Gandhi/Sun). The study was also supported in part by the following NIH/NIAID grants: K24AI114444 (PI: Gandhi), K24AI155045 (PI: Brust), K24AI165099 (PI: Shah); Emory/Georgia Tuberculosis Research Advancement Center (TRAC) P30AI168386 (MPIs: Gandhi/Rengarajan), Einstein-Rockefeller-CUNY Center for AIDS Research [grant P30AI124414], Einstein/Montefiore Institute for Clinical and Translational Research [grant UL1TR001073]).

### RePORT South Africa and HomeACF Acknowledgements

Participant recruitment and follow up was supported by the UK/South Africa Medical Research Council (MRC) Newton Fund (006Newton TB); and by the NIH through CRDF Global (DAA2-16-62117-1). The Free State, Limpopo and the North West Provincial Departments of Health facilitated and hosted these studies.

### RePORT India Acknowledgements

Participant recruitment and follow up was supported by Federal funds from the Government of India’s Department of Biotechnology (DBT) Project number BT/MB/COMMON PROTOCOL/01/2014-15 sanctioned to Bhagawan Mahavir Medical Research Centre (BMMRC), the Indian Council of Medical Research (ICMR), the United States NIH, NIAID, Office of AIDS Research (OAR), and distributed in part by CRDF Global.

RePORT India Participating Institutions, Principal investigators, co-principal investigators, DBT, ICMR, and NIAID staff include: Byramjee Jeejeebhoy Government Medical College (BJGMC): Aarti Kinikar, Sanjay Gaikwad, Rajesh Kayakarte; Byramjee Jeejeebhoy Government Medical College–Johns Hopkins India Clinical Research Site: Vidya Mave, Mandar Paradkar, Nikhil Gupte, Nishi Suryavanshi; National Institute for Research in Tuberculosis (NIRT): P. K. Bhavani, Elizabeth Hanna; Johns Hopkins University (JHU): Amita Gupta, Robert Bollinger, Jeff Tornheim; Bhagawan Mahavir Medical Research Centre (BMMRC): Vijaya Valluri; Saint Louis University: Ramakrishna Vankayalapati; Jawaharlal Institute of Postgraduate Medical Education and Research (JIPMER): Sonali Sarkar, Gautam Roy; Rutgers: Padmini Salgame, Jerrold Ellner; Centre for Cellular & Molecular Biology (CCMB): Vinay Nandicoori; DBT: Jyoti Logani; NIAID: Peter Kim, Fatima Jones; and CRDF Global: Tara Moore.

### RePORT-Brazil Acknowledgements

Data in this manuscript were collected as part of the RePORT-Brazil Consortium. This project has been funded in part with Federal funds from the Brazilian Ministry of Health (MoH), the National Council for Scientific and Technological Development (CNPq), the Fundação Oswaldo Cruz (Fiocruz), and the US NIH. It was also supported in part by NIH/NIAID grants: U01AI069923 (MPIs: Castillo, Cahn, Duda), R01AI120790 (MPIs: Sterling, Rolla), U01AI172064 (MPIs: Sterling, Andrade), U01AI174268 (MPIs: Ellner/Sterling/Andrade), R01AI147765 (MPIs: Sterling/Andrade/Hawn), AK, BBA, and MC-S are fellows from the Conselho Nacional de Desenvolvimento Científico e Tecnológico (CNPq), Brazil.

RePORT-Brazil Participating Institutions, Principal and Co-Investigators include:

Fundação José Silveira (FJS), Instituto Brasileiro para a Investigação da Tuberculose (IBIT), and Fundação Oswaldo Cruz–Bahia (Fiocruz-BA): Bruno B. Andrade;

Fundação de Medicina Tropical Dr. Heitor Vieira Dourado (FMT-HVD): Marcelo Cordeiro-Santos;

Instituto Nacional de Infectologia Evandro Chagas (INI–Fiocruz): Valeria Rolla;

Municipal Health Rinaldo De Lamare (Rocinha), Rio de Janeiro: Solange Cavalcante, Betina Durovni;

Municipal Health Department of Duque de Caxias, and Universidade Federal do Rio de Janeiro (UFRJ): Afranio Kritski;

Vanderbilt University Medical Center (VUMC): Timothy R. Sterling, Marina Cruvinel Figueiredo, and Megan Turner

The contents of this publication are solely the responsibility of the authors and do not necessarily represent the official views of the US NIH; South Africa MRC; India DBT or ICMR; Brazil MoH, CNPq, Fiocruz, or CRDF Global. Any mention of trade names, commercial products, or organizations does not imply endorsement by any of the sponsoring agencies.

## Supporting information

Full Supplemental Documentation

## Data Availability

All analytical code scripts are available online at the Sun Lab Git Hub page (https://github.com/Sun-Epi3-Lab/TB_GWAS.git).
Due to our ethics agreement, the analytic datasets used for this study require IRB approval. Thus, we share the genomic data and related phenotypes in NIH's dbGaP repository through controlled access. Full summary statistics of GWAS results will be available through GWAS catalog.

## Notes

### Competing Interest Statement

The authors have declared no competing interest.

### Author Declarations

IRB of Emory University, Johns Hopkins University, Vanderbilt University, Rutgers University, Comissão Nacional De Ética em Pesquisa (CONEP), University of Witswatersrand, Byramjee Jeejeebhoy Government Medical College, Jawaharlal Institute of Postgraduate Medical Education and Research, Bhagwan Mahavir Medical Research Centre, and the National Institute for Research in Tuberculosis (Chennai) gave ethical approval for this work.

