## Supplementary material for "Novel Genetic Locus Associated with Resistance to *M. tuberculosis* Infection: A Multi-Ancestry Genome-Wide Association Study": Full Supplemental Documentation

**Supplement**

### Supplemental Methods

#### Measurements of *Mtb* infection, index patient infectiousness, and exposure

IGRA results: We collated results of all interferon gamma release assays (IGRA) performed for each participant. Cohorts utilized the Quantiferon Gold-in-Tube (3^rd^ generation, Qiagen) or Quantiferon Gold Plus (4^th^ generation, Qiagen) assays. A threshold of 0.35 IU/ml was used to determine positivity. For IGRA-negative participants, the IGRA test was repeated at varying intervals (e.g., every 6 months), depending on the cohort. For persistently IGRA-negative participants, the number of days between the index patient’s diagnosis and the contact’s last IGRA test measured was determined; this measurement allowed us to assess if sufficient time had passed to allow for IGRA conversion as a result of this most recent TB exposure. Participants were categorized as: IGRA-positive; IGRA-negative (>90 days); IGRA-negative (<90 days).

TST results: We collated results of all tuberculin skin tests (TST) performed for each participant. A threshold of 5 mm of induration was used to determine positivity. For TST-negative participants, the TST was repeated at varying intervals, depending on the cohort. For persistently TST-negative participants, the number of days between the index patient’s diagnosis and the contact’s last TST test measured was determined; this measurement allowed us to assess if sufficient time had passed to allow for TST conversion. Participants were categorized as: TST-positive; TST-negative (>90 days); TST-negative (<90 days). TST were routinely performed only in cohorts from India. Difficulty importing tuberculin and issues related to standardization TST resulted in only a small minority of participants from Brazil and South Africa having TST results available.

At any study timepoint, if both an IGRA and TST were performed, the blood sample for IGRA was collected first; only after the IGRA sample was collected was the TST administered. At follow-up visits, TST or IGRA were only repeated if the prior visit test result for the respective test was negative.

Index patient infectiousness: Chest x-ray and AFB smear microscopy data of the participants’ index patients were collated to categorize infectiousness of the TB patients that participants were exposed to. Infectiousness was categorized as:

*High*: Cavities present on chest x-ray, or AFB smear positive with a grade of 2+ or 3+.

*Medium*: No cavities on chest x-ray and AFB smear positive with a grade scanty or 1+.

*Low*: No cavities on chest x-ray and AFB smear negative.

Exposure to the index case: Participants exposure to the TB index patient was characterized based on whether they slept in the same bed or room as the index patient and the number of hours spent indoors with the patient.

*High*: Participant slept in same bed or room, or spent ≥5 hours indoors with the index patient before their TB diagnosis.

*Low*: Participant did not sleep in the same room and spent <5 hours indoors with the index patient.

#### Characterization of Resister Phenotype

We separated participants into categories based on the data available regarding *Mtb* infection status, infectiousness of index patients, and exposure to index patient. The goal of these categories was to create strata which reflect the likelihood that the participants were resistant to *Mtb* infection, or the likelihood they were *Mtb* infected (**Figure 1**).

Uninfected categories: Participants whose results for all IGRA or TST tests performed were negative were classified into 5 categories: Resister A/B/C and Uninfected A-B. Participants who were exposed to an index patient with “high” infectiousness (e.g., cavities, smear 2+/3+) and had a “high” degree of exposure (e.g., slept in same room) were placed into categories Resister A/B/C.

*Resister A*: Highly exposed participants who had **both** a negative IGRA and negative TST and at least one of these tests was performed ≥90 days after the index patient’s diagnosis.

*Resister B*: Highly exposed participants with either only IGRA result(s) or TST result(s), but not both; the latest of those results was ≥90 days after index case diagnosis.

*Resister C*: Highly exposed participants with IGRAs or TSTs, whose latest of those tests is <90 days after index patient diagnosis.

*Uninfected A*: Uninfected participants whose index patient’s infectiousness was medium or low, or whose exposure to the index patient was low.

*Uninfected B*: Uninfected participants who were missing either index patient’s infectiousness data or exposure data.

Infected categories: Participants whose results for all IGRA and TST tests were positive were separated into two categories: Infected A & B.

*Infected A* had both IGRA and TST results available.

*Infected B* had either an IGRA result available or a TST result, but not both.

Discordant category: Participants who had both IGRA and TST results available, with at least one result positive and one result negative.

#### Sensitivity Analyses for GWAS Analyses

For the GWAS for resistance, we conducted sensitivity analyses by redefining both the control groups and the resister group.

Sensitivity Analysis #1: We repeated the GWAS using only participants classified as Infected A–B or Discordant as the control group, thereby excluding individuals in the Uninfected A-B categories from the analysis (**Supplemental Table S7**). This approach was intended to minimize potential misclassification of uninfected individuals as not resistant, since their resistance status was less certain because of either missing data or low exposure.

Sensitivity Analysis #2: We tested an alternative definition of resisters by including participants categorized as Uninfected B, who had either exposure or infectiousness data missing, alongside those in Resister A–C. This sensitivity analysis allowed for the inclusion of individuals who may have had high degree of exposure or infectiousness, if their data were available. Since a large proportion of participants were missing data on index case infectiousness, this analysis allowed us to perform the GWAS for resistance for participants from the South African cohorts.

Both sensitivity analyses allowed us to evaluate how findings would differ if the resister phenotype were expanded beyond the stringent definition used in the main analysis.

For the GWAS for *Mtb* infection, we examined the effect of broadening the case definition to incorporate participants with discordant TST and IGRA results.

Sensitivity Analysis #3: The Discordant group was combined with Infected A–B to create an expanded case set, which was then contrasted with all other participants. This analysis tested whether inclusion of individuals who may be potentially misclassified as *Mtb* infected influenced effect estimates or altered the loci of interest identified in the primary analysis.

Quality control, imputation, and covariate adjustment procedures were identical to those used in the main GWAS.

#### Ethical Considerations

This study was approved by the institutional review boards at the following institutions: Emory University, Johns Hopkins University, Vanderbilt University, Rutgers University, Commisão Nacional De Ética em Pesquisa (CONEP), University of Witswatersrand, Byramjee Jeejeebhoy Government Medical College, Jawaharlal Institute of Postgraduate Medical Education and Research, Bhagwan Mahavir Medical Research Centre, and National Institute for Research in Tuberculosis (Chennai).

### Supplemental Results

#### Sensitivity Analyses #1 and #2 for GWAS for resistance

The GWAS for resistance sensitivity analysis #1, restricting the control group to Infected A–B and Discordant individuals (n = 448 cases, n = 1,847 controls, **Supplemental Table S7a**), yielded results highly consistent with those reported in the primary analysis. The lead SNP at chromosome 13 (rs1295104126) remained genome-wide significant with a similar effect size, though the precision of the estimate was slightly reduced (**Supplemental Figure S6a)**. While other top SNPs identified in the main analysis remained directionally similar, some associations showed attenuation in significance, likely reflecting the reduced number of controls in this restricted comparison (from 2,710 to 1,847).

In Sensitivity Analysis #2, when the resister group was expanded to include Uninfected B participants (n = 957 cases, n = 2,932 controls), the effect estimate at the same chromosome 13 locus diminished substantially and no longer reached genome-wide significance (**Supplemental Figure S6a)**. Nevertheless, the SNPs with the highest associations under this broader definition are still located within the same chromosome 13 locus identified in our main analysis, suggesting that the association was not spurious but sensitive to alterations to the phenotype definition.

Overall, these results demonstrate that excluding individuals of uncertain resistance status (Uninfected A-B) from the control group had minimal impact. However, including participants with missing exposure or infectiousness data (Uninfected B) in the resister group introduces considerable heterogeneity that weakens the originally observed association signals.

#### Sensitivity Analyses #3 for GWAS for Mtb infection

For the GWAS for *Mtb* infection Sensitivity Analysis #3, we redefined the case group to include Discordant individuals in addition to those who are Infected A and B (n = 2,274 cases, n = 1,615 controls; **Supplemental Table S7b**). Under this broader case definition, the primary chromosome 6 association (SNP rs28752534) remained genome-wide significant but with a weaker effect size and wider confidence intervals compared to the results from the main analysis (**Supplemental Figure S6b**). These findings suggest that the inclusion of individuals with discordant TST and IGRA results only slightly alters the original findings, but the observed genetic architecture for infection susceptibility remained the same.

Through these sensitivity analyses we have demonstrated that the main associations at chromosome 13 and chromosome 6 were reproducible across phenotype definitions. As expected, the precision and effect sizes were impacted differently depending on the inclusions and exclusions applied, but the overall genomic signals remained stable.

### Supplemental Tables & Figures

#### Supplemental Table S1: Number of participants enrolled from each of the collaborating cohorts

| Country | Collaborating Institution | Cohort | N enrolled | % enrolled |
| --- | --- | --- | --- | --- |
| Brazil | Oswaldo Cruz Foundation (Fiocruz) – Instituto Nacional de Infectologia (INI) – Rio de Janeiro | RePORT Common Protocol | 1558 | 38% |
|  | Rio de Janeiro Municipal Health Secretariat – Rocinha – Rio de Janeiro |  |  |  |
|  | Centro Municipal de Saúde de Duque de Caxias and UFRJ – Rio de Janeiro |  |  |  |
|  | Fundação de Medicina Tropical (FMT) – Manuas |  |  |  |
|  | Instituto Brasileiro para Investigação da Tuberculose (IBIT) – Salvador, Bahia |  |  |  |
| India | Byramjee Jeejeebhoy Government Medical College (BJGMC) – Pune | RePORT Common Protocol | 213 | 5.2% |
|  |  | RePORT Parent Protocol | 349 | 8.6% |
|  |  | TB GWAS Prospective cohort | 201 | 5.0% |
|  | Jawaharlal Institute of Postgraduate Medical Education and Research (JIPMER) – Puducherry | RePORT Parent Protocol | 555 | 14% |
|  |  | RePORT Common Protocol | 187 | 4.6% |
|  | Bhagwan Mahavir Medical Research Centre (BMMRC) – Hyderabad | RePORT Common Protocol | 153 | 3.8% |
| South Africa | Perinatal HIV Research Unit (PHRU) – Matlosana | RePORT SoHOT cohort | 198 | 4.9% |
|  | Perinatal HIV Research Unit (PHRU) – Botshabelo | Home ACF cohort | 644 | 16% |
| Total |  |  | 4058 |  |

#### Supplemental Table S2: Description of participants, exposure characteristics, and IGRA and TST results by cohort and country

|  | **Total**  **(N=4058)** | **Brazil Cohorts**  **(N=1558)** | **BJGMC CP (N=213)** | **BJGMC PP (n=349)** | **BJGMC Prospective (n=201)** | **JIPMER CP (n=187)** | **JIPMER PP (n=555)** | **BMMRC CP (n=153)** | **India Total**  **(N=1658)** | **SOHOT (n=198)** | **Home ACF (n=644)** | **South Africa Cohorts**  **(N=842)** |
| --- | --- | --- | --- | --- | --- | --- | --- | --- | --- | --- | --- | --- |
| **Sex: Female** | 2385 (59%) | 934 (60%) | 109 (51%) | 199 (57%) | 100 (50%) | 128 (68%) | 314 (57%) | 87 (57%) | 937 (57%) | 116 (59%) | 398 (62%) | 514 (61%) |
| **Age: median years (IQR)** | 27 (15-43) | 32 (16-47) | 27 (16-40) | 25 (12-37) | 32 (22-40) | 28 (17-43) | 24 (15-38) | 30 (20-38) | 26 (16-39) | 24 (14-47) | 18 (12-38) | 20 (12-39) |
| **<5** | 98 (2.4%) | 66 (4.2%) | 10 (4.7%) | 17 (4.9%) | 0 (0.0%) | 0 (0.0%) | 0 (0.0%) | 0 (0.0%) | 27 (1.6%) | 0 (0.0%) | 5 (0.8%) | 5 (0.6%) |
| **5-12** | 637 (16%) | 212 (14%) | 29 (14%) | 71 (20%) | 1 (0.5%) | 24 (13%) | 82 (15%) | 2 (1.3%) | 209 (13%) | 31 (16%) | 185 (29%) | 216 (26%) |
| **13-20** | 816 (20%) | 222 (14%) | 35 (16%) | 62 (18%) | 44 (22%) | 46 (25%) | 154 (28%) | 37 (24%) | 378 (23%) | 43 (22%) | 173 (27%) | 216 (26%) |
| **21-30** | 719 (18%) | 228 (15%) | 52 (24%) | 59 (17%) | 50 (25%) | 26 (14%) | 116 (21%) | 46 (30%) | 349 (21%) | 49 (25%) | 93 (14%) | 142 (17%) |
| **31-40** | 666 (16%) | 274 (18%) | 35 (16%) | 77 (22%) | 56 (28%) | 36 (19%) | 85 (15%) | 44 (29%) | 333 (20%) | 21 (11%) | 38 (5.9%) | 59 (7.0%) |
| **41-50** | 498 (12%) | 236 (15%) | 34 (16%) | 40 (12%) | 28 (14%) | 28 (15%) | 70 (13%) | 17 (11%) | 217 (13%) | 14 (7.1%) | 31 (4.8%) | 45 (5.3%) |
| **51-60** | 348 (8.6%) | 187 (12%) | 15 (7%) | 14 (4.0%) | 16 (8.0%) | 16 (8.6%) | 36 (6.5%) | 7 (4.6%) | 104 (6.3%) | 17 (8.6%) | 40 (6.2%) | 57 (6.8%) |
| **61-70** | 191 (4.7%) | 100 (6.4%) | 3 (1.4%) | 9 (2.6%) | 4 (2.0%) | 9 (4.8%) | 7 (1.3%) | 0 (0.0%) | 32 (1.9%) | 15 (7.6%) | 44 (6.8%) | 59 (7.0%) |
| **>70** | 85 (2.1%) | 33 (2.1%) | 0 (0.0%) | 0 (0.0%) | 2 (1.0%) | 2 (1.1%) | 5 (0.9%) | 0 (0.0%) | 9 (0.5%) | 8 (4.0%) | 35 (5.4%) | 43 (5.1%) |
| **Exposure to Index patient** |  |  |  |  |  |  |  |  |  |  |  |  |
| **Slept in same bed** | 554 (17%) | 182 (18%) | 51 (30%) | 75 (22%) | 54 (27%) | 15 (7.6%) | 42 (7.6%) | - | 237 (16%) | 26 (15%) | 109 (17%) | 135 (17%) |
| **Slept in same room** | 860 (26%) | 156 (15%) | 73 (44%) | 164 (47%) | 84 (42%) | 95 (51%) | 201 (36%) | - | 617 (42%) | 13 (7.6%) | 74 (12%) | 87 (11%) |
| **Spent ≥5 hours indoors together** | 1230 (37%) | 542 (52%) | 43 (26%) | 86 (25%) | 57 (28%) | 53 (28%) | 0 (0.0%) | - | 239 (16%) | 0 | 449 (70%) | 449 (55%) |
| **None of the above** | 666 (20%) | 154 (15%) | 1 (0.6%) | 24 (6.9%) | 6 (3.0%) | 24 (13%) | 312 (56%) | - | 367 (25%) | 133 (77%) | 12 (1.9%) | 145 (18%) |
| **Not available** | 748 | 524 | 45 | 0 | 0 | 0 | 0 | 153 | 198 | 26 | 0 | 26 |
| **Index case infectiousness** |  |  |  |  |  |  |  |  |  |  |  |  |
| **High: Cavities present or AFB smear 2+/3+** | 2261 (64%) | 972 (62%) | 197 (92%) | 198 (57%) | 91 (47%) | 167 (89%) | 410 (74%) | 73 (48%) | 1136 (69%) | 119 (60%) | 34 (72%) | 153 (62%) |
| **Medium: AFB smear scanty/1+, OR  GeneXpert grade high or very high (no cavities)** | 776 (22%) | 324 (21%) | 2 (0.9%) | 93 (27%) | 91 (47%) | 20 (11%) | 142 (26%) | 80 (52%) | 428 (26%) | 11 (5.6%) | 13 (28%) | 24 (9.7%) |
| **Low: AFB smear negative OR**  **GeneXpert grade low or very low (no cavities)** | 416 (14%) | 262 (17%) | 14 (6.6%) | 58 (17%) | 11 (5.7%) | 0 (0.0%) | 3 (0.5%) | 0 | 86 (5.2%) | 68 (34%) | 0 | 68 (28%) |
| **Not available** | 605 | 0 | 0 | 0 | 8 | 0 | 0 | 0 | 8 | 0 | 597 | 597 |
| **IGRA results** |  |  |  |  |  |  |  |  |  |  |  |  |
| **Positive** | 1896 (55%) | 746 (48%) | 151 (72%) | 217 (63%) | 127 (65%) | 112 (62%) | 0 | 101 (66%) | 708 (65%) | 122 (66%) | 330 (54%) | 442 (56%) |
| **Negative (>90 days)** | 1311 (38%) | 794 (51%) | 55 (26%) | 80 (23%) | 51 (26%) | 34 (19%) | 0 | 0 | 220 (20%) | 13 (7.6%) | 284 (46%) | 297 (38%) |
| **Negative (<90 days)** | 221 (6.5%) | 16 (1.0%) | 5 (2.4%) | 49 (14%) | 18 (9.2%) | 36 (20%) | 0 | 52 (34%) | 160 (15%) | 45 (27%) | 0 (0.0%) | 45 (5.9%) |
| **Not available** | 630 | 2 | 2 | 3 | 5 | 5 | 555 | 0 | 570 | 28 | 30 | 58 |
| **TST results** |  |  |  |  |  |  |  |  |  |  |  |  |
| **Positive** | 805 (54%) | - | 76 (68%) | 182 (54%) | 123 (62%) | 55 (41%) | 277 (50%) | 92 (61%) | 805 (54%) | - | - | - |
| **Negative (>90 days)** | 154 (10%) | - | 10 (8.9%) | 91 (27%) | 52 (26%) | 0 | 1 (0.2%) | 0 | 154 (10%) | - | - | - |
| **Negative (<90 days)** | 529 (36%) | - | 26 (23%) | 66 (20%) | 22 (11%) | 78 (59%) | 277 (50%) | 60 (41%) | 529 (36%) | - | - | - |
| **Not available** | 2570 | 1558 | 101 | 10 | 4 | 54 | 0 | 1 | 170 | 198 | 644 | 842 |

CP=Common Protocol, PP=Parent Protocol, IGRA=interferon gamma release assay, TST=tuberculin skin test

#### Supplemental Figure S1: Flow diagram of participants enrolled by country and their inclusion in final GWAS analyses

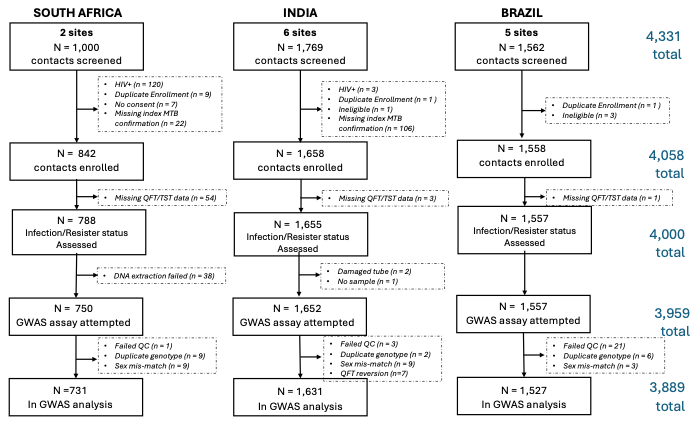

##### Supplemental Table S3: Consolidated phenotypic categories for use in GWAS analyses

| **Hierarchy** |  | **Total**  **N=4000**  **N (%)** | **Brazil**  **N=1557**  **N (%)** | **India**  **N=1655**  **N (%)** | **South Africa**  **N=788**  **N (%)** |
| --- | --- | --- | --- | --- | --- |
| Resistant to *Mtb* Infection | Resisters A/B/C | 476 (12%) | 231 (15%) | 227 (14%) | 18 (2.3%) |
| Not infected, unclear if resistant | Uninfected A-B | 1176 (29%) | 559 (36%) | 315 (19%) | 302 (38%) |
| Discordant infection results | Discordant | 357 (8.9%) | 7 (0.4%) | 329 (20%) | 21 (2.7%) |
| *Mtb* Infected | Infected A-B | 1991 (50%) | 760 (49%) | 784 (47%) | 447 (57%) |

#### Supplemental Table S4: Availability of data elements by collaborating cohort

|  | **Brazil** | **India**  **BJGMC  CP** | **India**  **BJGMC PP** | **India**  **BJGMC Prosp.** | **India**  **JIPMER CP** | **India**  **JIPMER  PP** | **India**  **BMMRC CP** | **South Africa**  **SoHOT** | **South Africa Home ACF** |
| --- | --- | --- | --- | --- | --- | --- | --- | --- | --- |
| **Exposure**  **data** | X | X | X | X | X | X |  | X | X |
| **Index infectiousness** | X | X | X | X | X |  | X | X |  |
| **QFT results** | X | X | X | X | X |  | X | X | X |
| **TST results** |  | X | X | X | X | X | X |  |  |

##### Supplemental Table S5: Comparison of demographics, exposure characteristics, and IGRA and TST results between Resister and other categories

|  | **Resisters A/B/C** | **Uninfected A-B** | **Discordant** | **Infected A-B** |
| --- | --- | --- | --- | --- |
| Age, median (IQR) | 28 (14-42) | 22 (12-38) | 28 (18-40) | 30 (17-46) |
| Sex, female | 284 (60%) | 663 (56%) | 195 (55%) | 1207 (61%) |
| IGRA results |  |  |  |  |
| Positive | 0 (0%) | 0 (0%) | 207 (58%) | 1689 (99%) |
| Negative (>90 days) | 338 (86%) | 879 (90%) | 82 (23%) | 12 (0.7%)* |
| Negative (<90 days) | 56 (14%) | 95 (9.8%) | 68 (19%) | 2 (0.1%)* |
| Not available | 82 | 202 | 0 | 288 |
| TST results |  |  |  |  |
| Positive | 0 (0%) | 0 (0%) | 150 (42%) | 843 (100%) |
| Negative (>90 days) | 40 (23%) | 51 (17%) | 63 (18%) | 0 (0%) |
| Negative (<90 days) | 132 (77%) | 253 (83%) | 144 (40%) | 0 (0%) |
| Not available | 304 | 872 | 0 | 1148 |
| Exposure to index case |  |  |  |  |
| Slept in same bed | 96 (20%) | 109 (12%) | 56 (19%) | 286 (18%) |
| Slept in same room | 175 (37%) | 141 (15%) | 129 (45%) | 410 (26%) |
| Spent ≥5 hours indoors together | 205 (43%) | 332 (36%) | 80 (28%) | 591 (37%) |
| None of the above | 0 (0%) | 338 (37%) | 23 (8.0%) | 284 (18%) |
| Not available | 0 | 256 | 69 | 420 |
| Index case infectiousness |  |  |  |  |
| **High:** Cavities present or AFB smear 2+/3+ | 476 (100%) | 357 (39%) | 207 (62%) | 1197 (71%) |
| **Medium:** AFB smear scanty/1+, OR  GeneXpert grade high or very high (no cavities) | 0 (0%) | 342 (37%) | 107 (32%) | 326 (19%) |
| **Low:** AFB smear negative OR  **G**eneXpert grade low or very low (no cavities) | 0 (0%) | 229 (25%) | 22 (6.5%) | 156 (9.3%) |
| Not available | 0 | 248 | 21 | 312 |
| Country of Enrollment |  |  |  |  |
| South Africa | 18 (3.8%) | 302 (26%) | 21 (5.9%) | 447 (23%) |
| India | 227 (48%) | 315 (27%) | 329 (92%) | 784 (39%) |
| Brazil | 231 (49%) | 559 (48%) | 7 (2.0%) | 760 (38%) |

**Individuals categorized as “infected” based on a previous history of active TB disease, despite having negative IGRA results in our study*

Supplemental Figure S2: Principal Component Analysis plots for participants from India, South Africa, and Brazil: red triangles indicate study participants; dots represent reference samples of African (black dots), East Asian (dark green), European (blue), and South Asian (light green) ancestry.

**Supplemental Figure S2a. Participants from Indian cohorts:** Participants’ PCA plots (red triangles) overlap with reference samples of South Asian ancestry (light green dots)

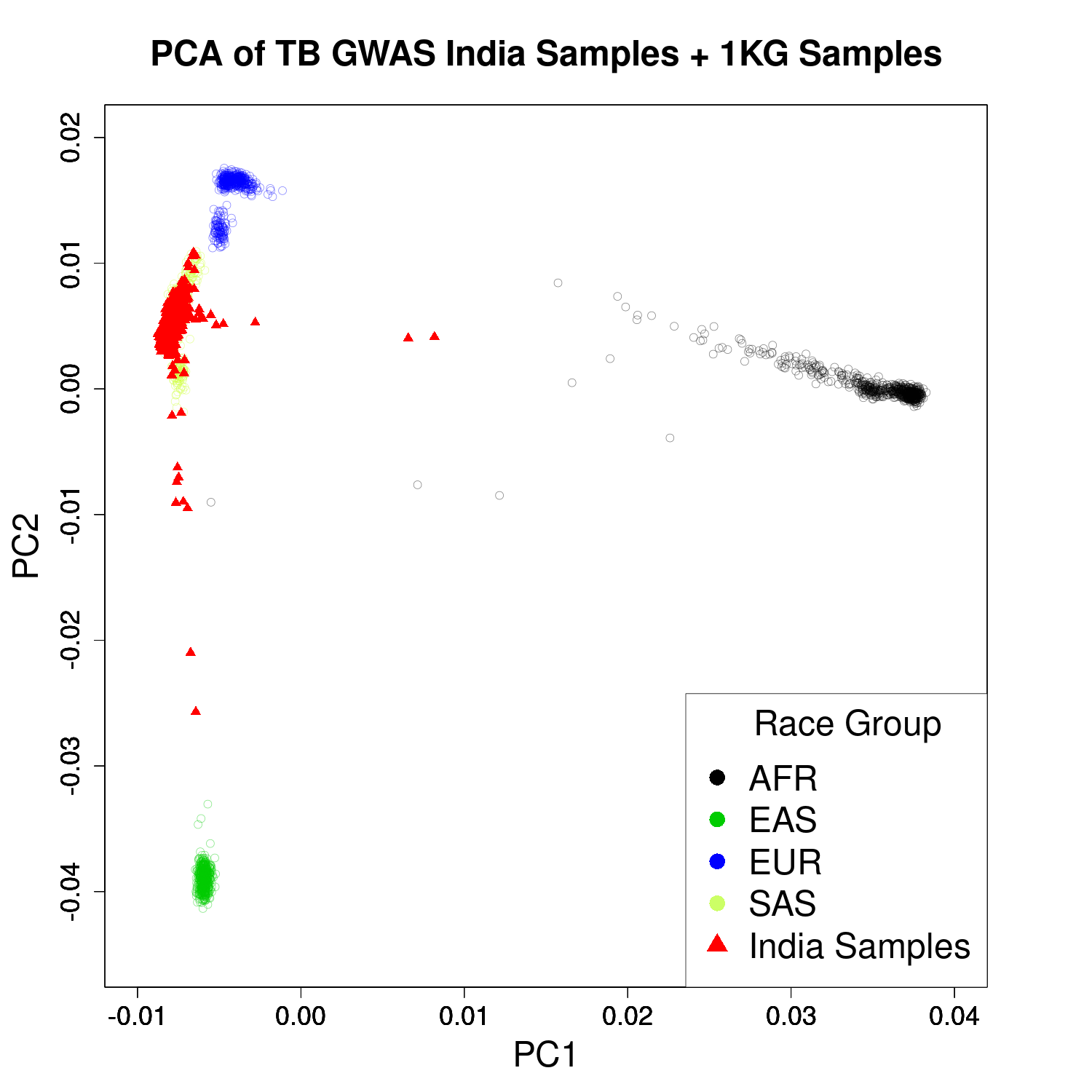

**Supplemental Figure S2b. Participants from South African cohorts:** Participants’ PCA plots (red triangles) overlap with reference samples of African ancestry (black dots)

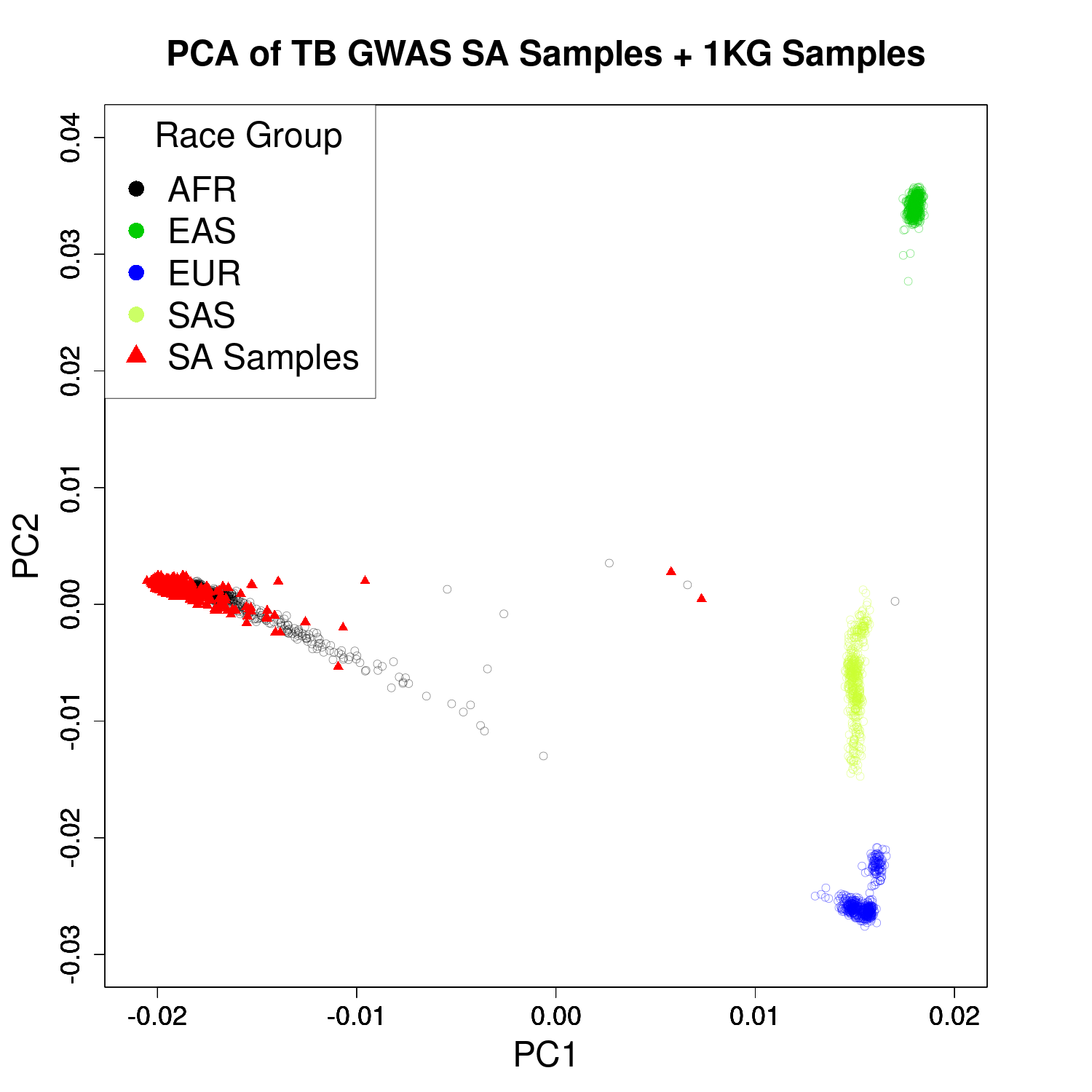

**Supplemental Figure S2c. Participants from Brazilian cohorts:** Participants’ PCA plots (red triangles) show admixture of ancestries between reference samples of African (black dots), European (blue dots), and South Asian (light green dots) ancestry.

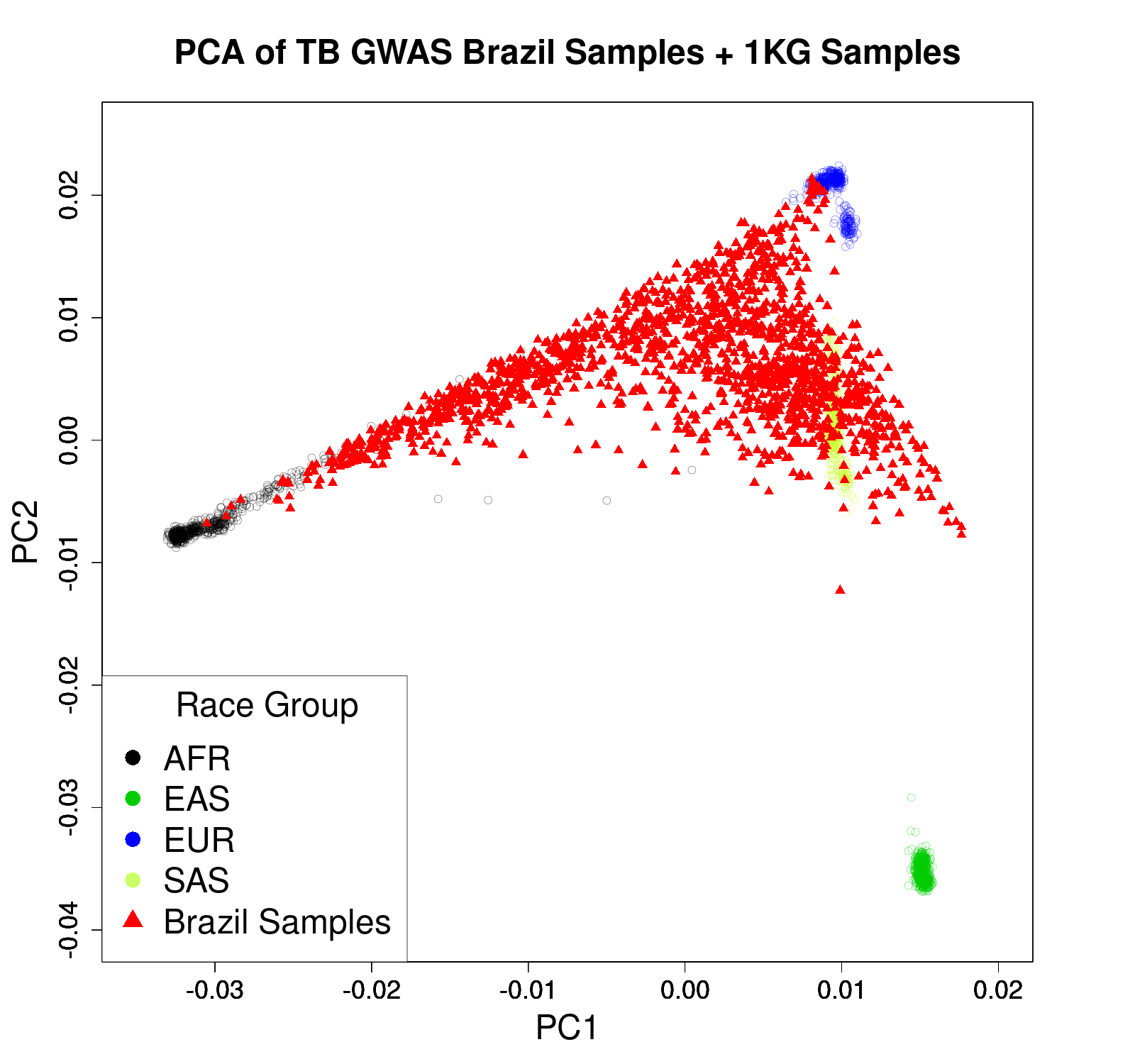

#### Supplemental Figure S3: Genetic markers from GWAS for resistance by individual countries: India, Brazil

**Supplemental Figure S3a. India**: One statistically significant association found on chromosome 13 (SNP rs11843280, OR = 2.53, 95% CI 1.83 – 3.50, p = 1.71x10^-8^)

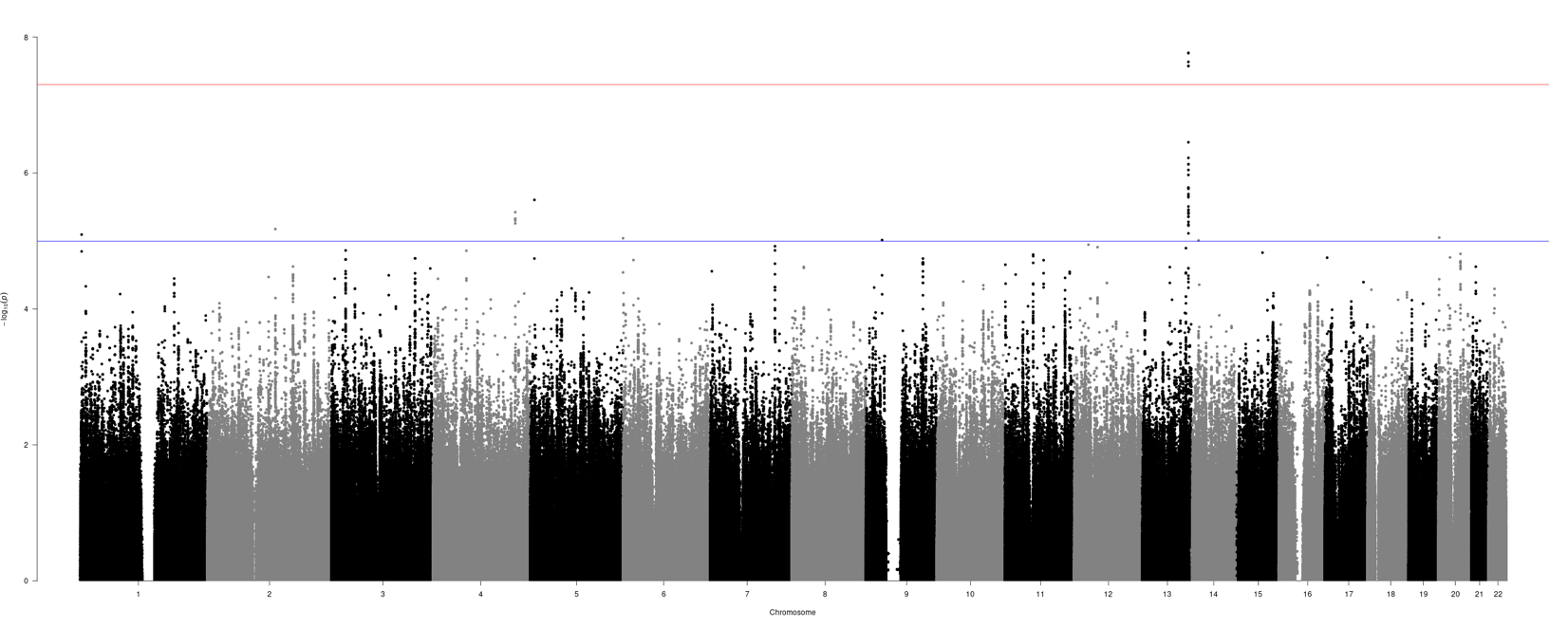

**Supplemental Figure S3b. Brazil:** No statistically significant loci identified at the genome-wide threshold of p<5x10^-8^. The significant locus from the India GWAS (SNP rs11843280) had the same direction of effect but did not achieve the GWAS threshold of statistical significance (OR = 1.74, 95%CI 1.26 – 2.40, p = 0.0007).

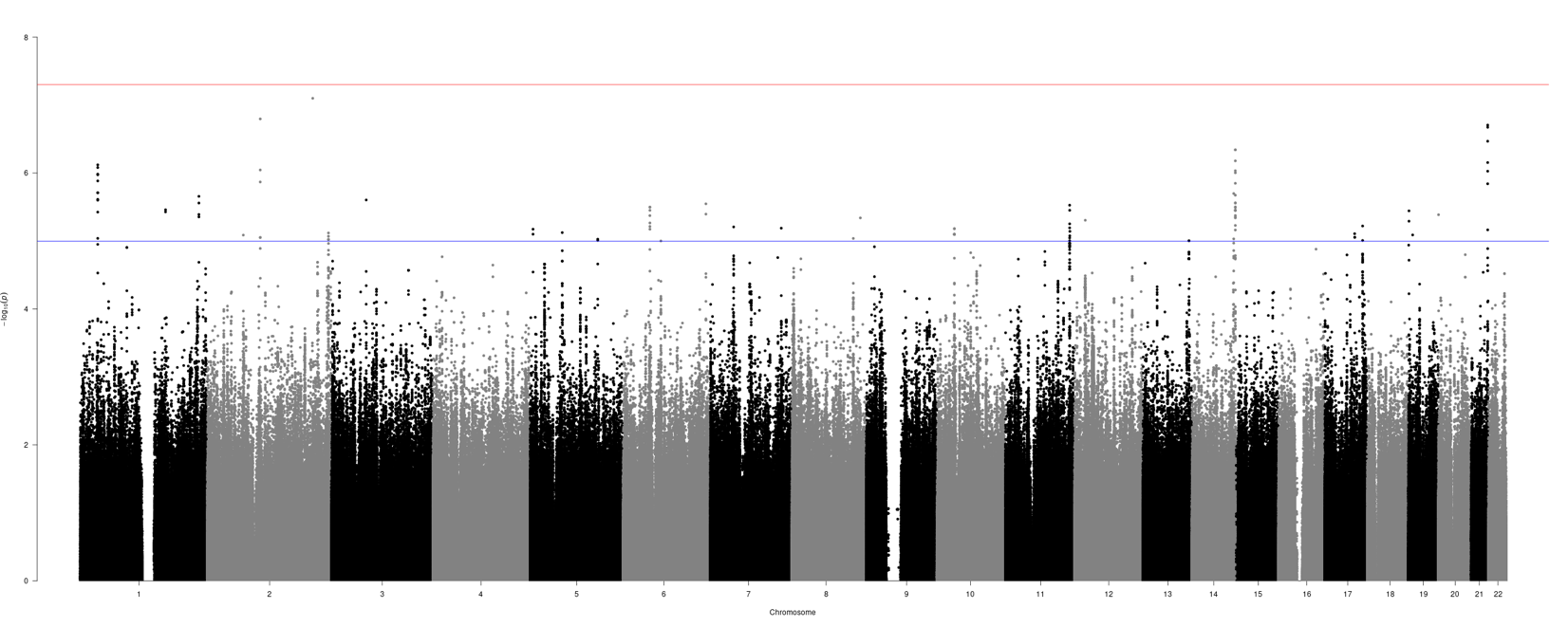

#### Supplemental Table S6: Tables of statistically significant Single Nucleotide Polymorphisms (SNPs) from the GWAS for resistance and GWAS for *Mtb* infection meta-analyses and individual country analyses

**Supplemental Table S6a**. GWAS for resistance SNP results from meta-analysis and Brazil and India country analyses

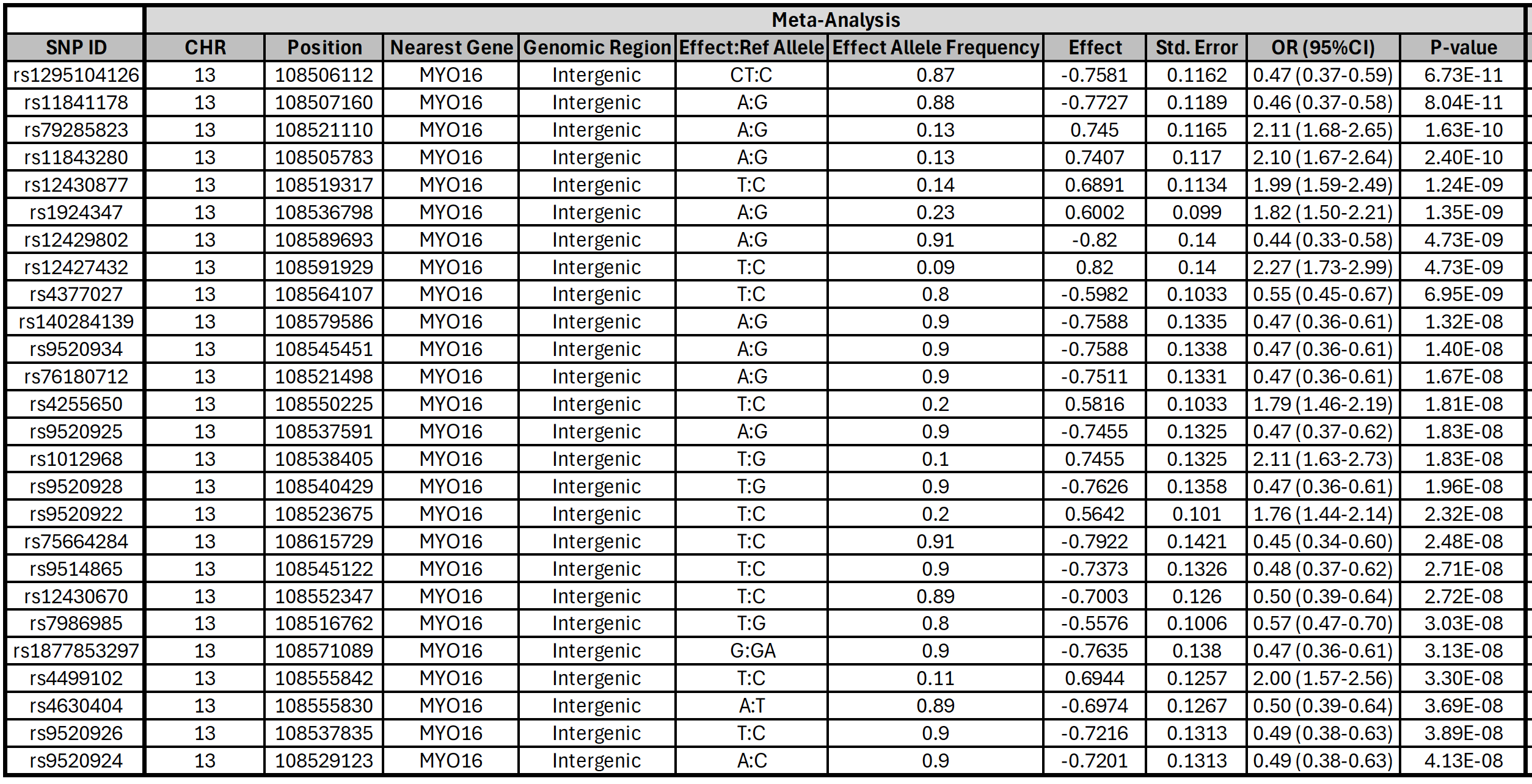

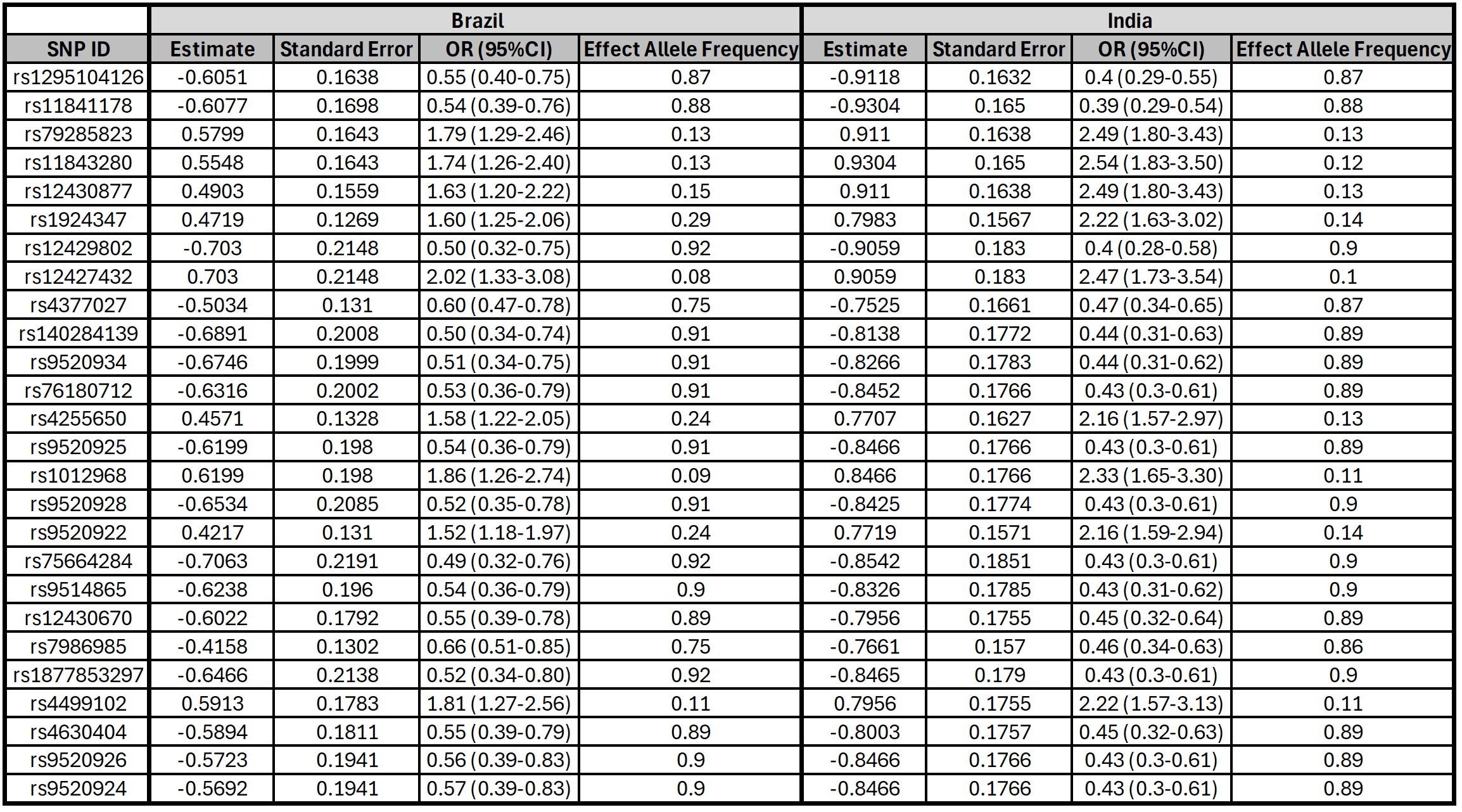

**Supplemental Table S6b**: GWAS for *Mtb* infection SNP results from meta-analysis and Brazil and India country analyses

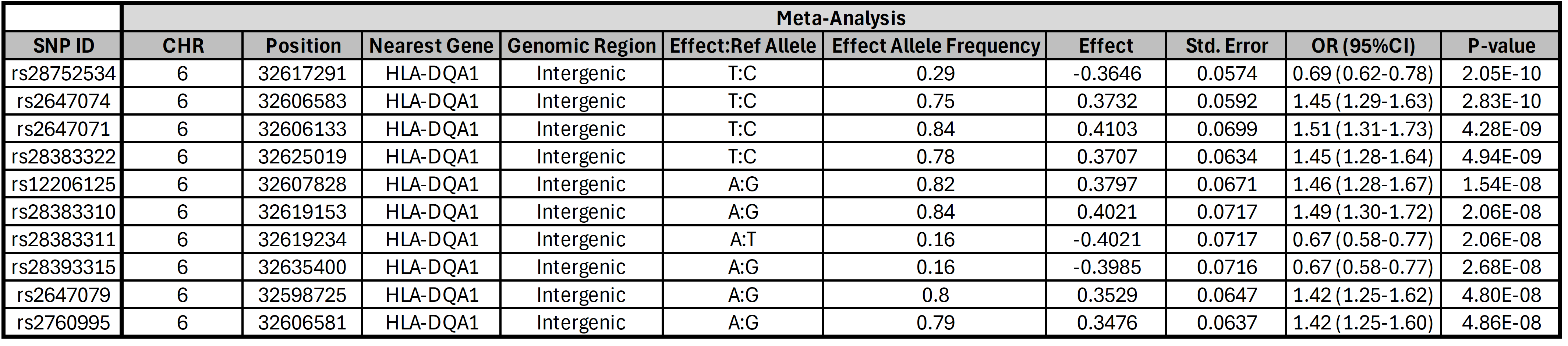

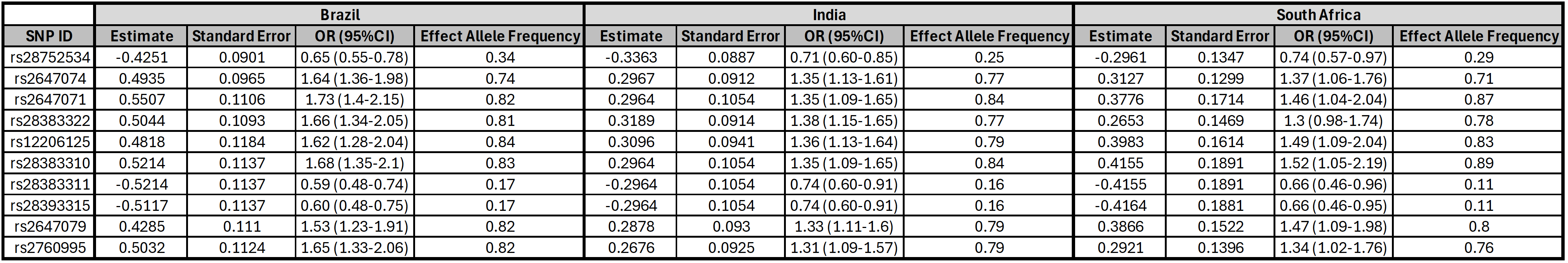

#### Supplemental Figure S4: Forest Plots of GWAS for resistance and GWAS for *Mtb* infection by country and meta-analysis

**Supplemental Figure S4a.** Forest Plots of GWAS for resistance by country and meta-analysis for SNP rs1295104126

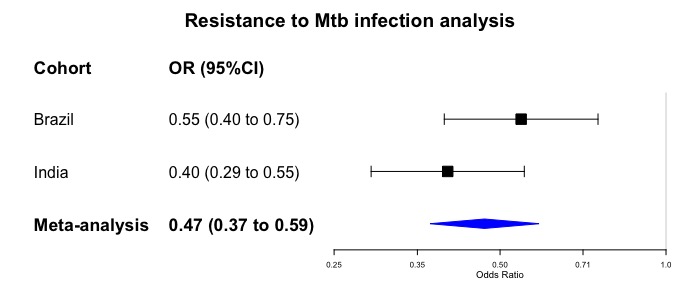

**Supplemental Figure S4b.** Forest Plots of GWAS for *Mtb* infection by country and meta-analysis for SNP rs28752534

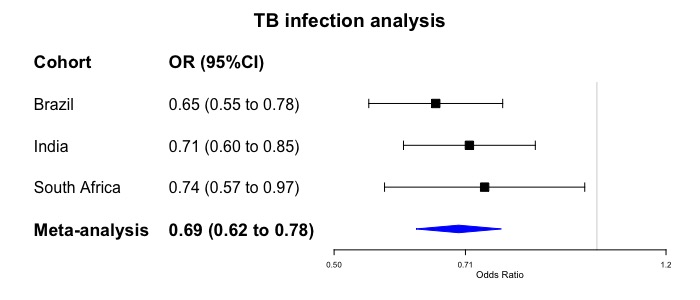

#### Supplemental Figure S5: Quantile-quantile (Q-Q) plots for GWAS for resistance and GWAS for *Mtb* infection analyses

**Supplemental Figure 5a.** Quantile-quantile (Q-Q) plot for GWAS for resistance meta-analysis (India and Brazil cohorts only): Inflation factor is 1.005**.**

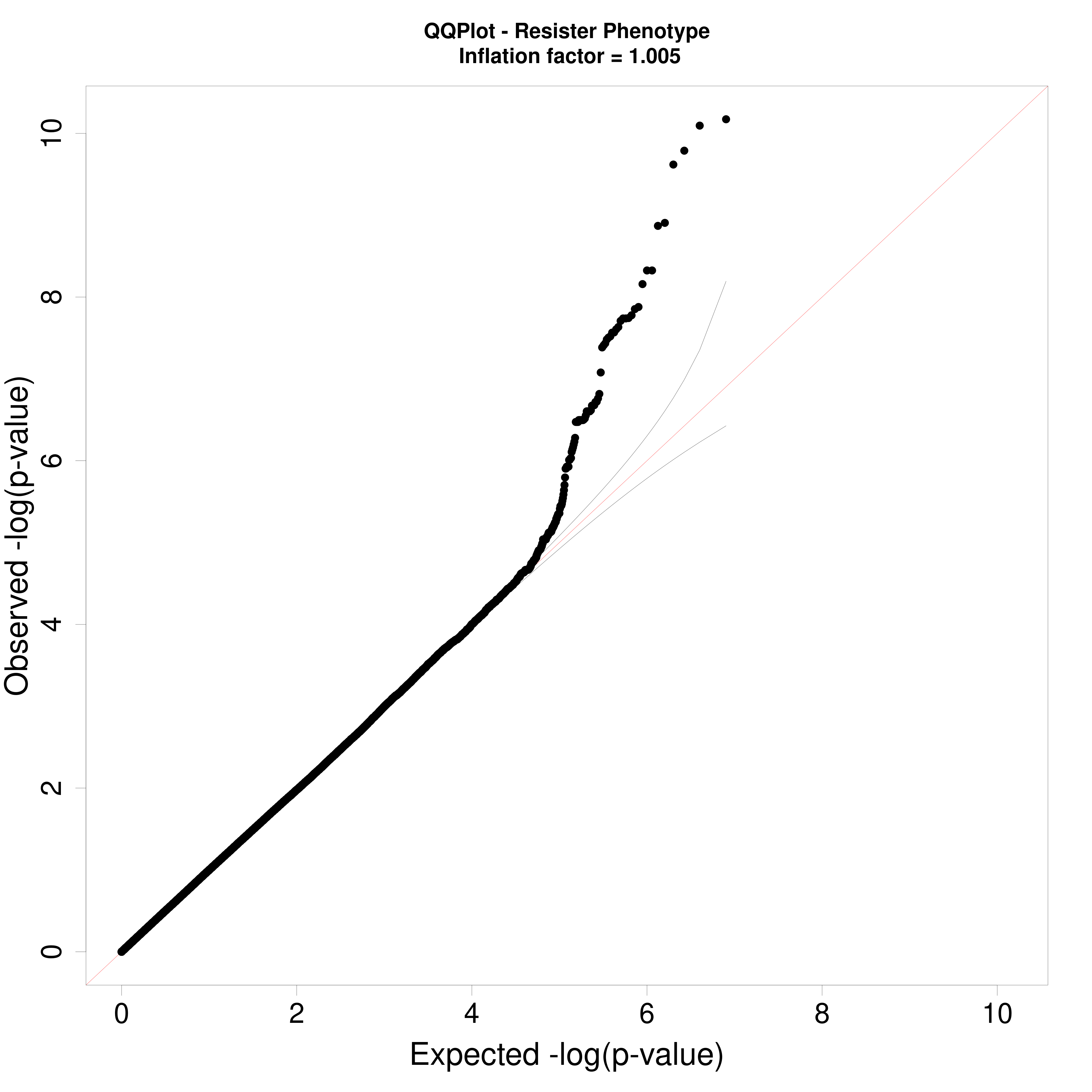

**Supplemental Figure S5b.** Quantile-quantile (Q-Q) plot for GWAS for *Mtb* Infection (meta-analysis (India, Brazil, and South Africa cohorts): Inflation factor is 1.001.

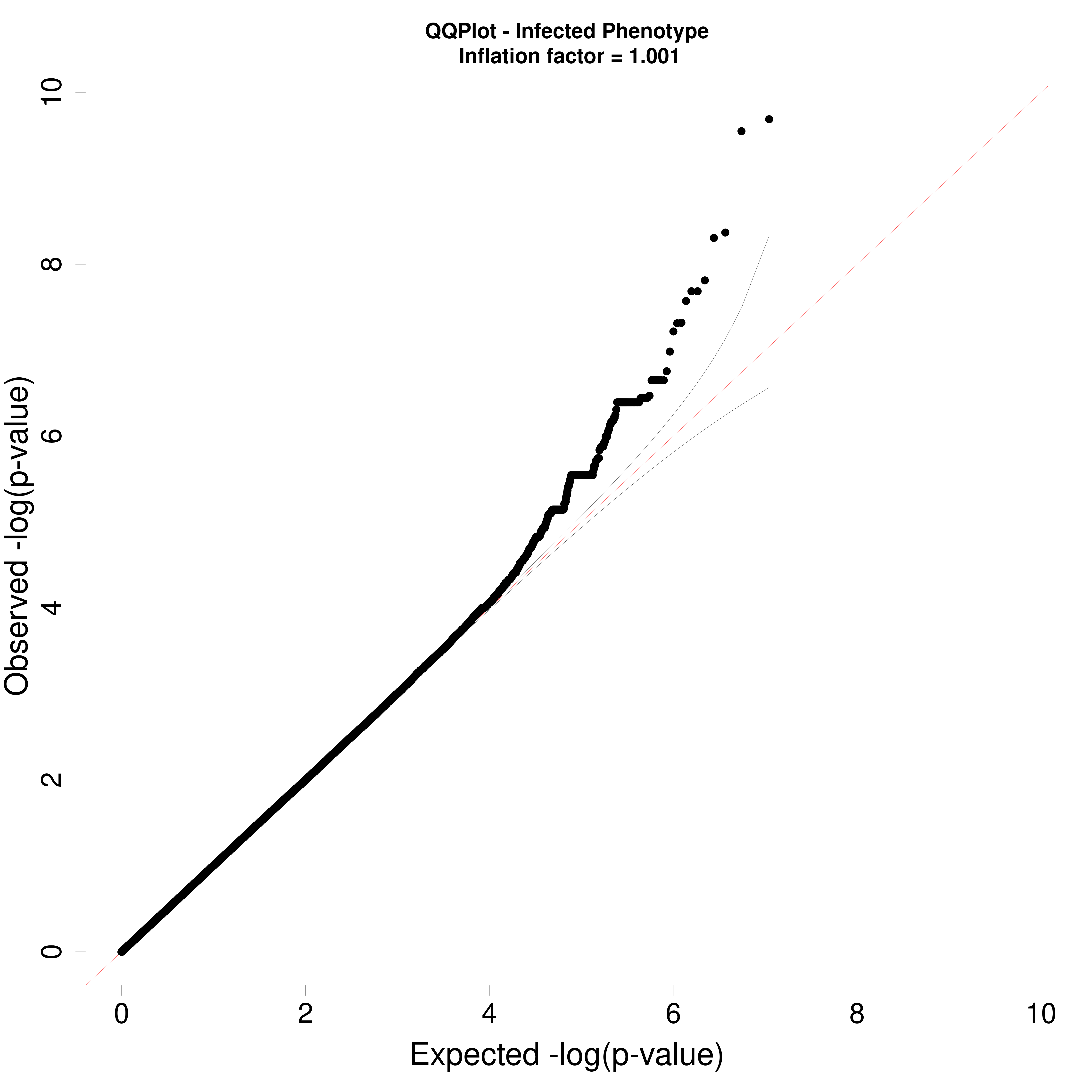

#### Supplemental Table S7: Comparison groups in each of the main GWAS analyses and sensitivity analyses

**Supplemental Table S7a**: Comparison groups for GWAS for resistance and Sensitivity Analyses #1 & #2, varying the resister case and control groups

| **Hierarchy** | **Main Resistance**  **GWAS analysis**  (Only India & Brazil) | **Resister Sensitivity Analysis #1**  (Only India & Brazil) | **Resister Sensitivity Analysis #2**  (India, Brazil, South Africa) |
| --- | --- | --- | --- |
| Resister A | Resisters  (N = 448) | Resisters  (N = 448) | Resisters (N=957) |
| Resister B |  |  |  |
| Resister C |  |  |  |
| Uninfected A | Controls  (N= 2710) | Excluded | Controls  (N= 2932) |
| Uninfected B |  |  | Resisters (N=957) |
| Discordant |  | Controls  (N= 1847) | Controls  (N= 2932) |
| Infected B |  |  |  |
| Infected A |  |  |  |

**Supplemental Table S7b:** Comparison groups for GWAS for *Mtb* infection and Sensitivity Analysis #3, varying the infected cases of interest

| **Hierarchy** | **Main *Mtb* Infection**  **GWAS analysis**  (India, Brazil, South Africa) | ***Mtb* Infection**  **Sensitivity Analysis #3** (India, Brazil, South Africa) |
| --- | --- | --- |
| Resister A | Controls (N=1967) | Controls  (N = 1615) |
| Resister B |  |  |
| Resister C |  |  |
| Uninfected A |  |  |
| Uninfected B |  |  |
| Discordant |  | Infected  (N= 2274) |
| Infected B | Infected (N=1922) |  |
| Infected A |  |  |

#### Supplemental Figure S6: Sensitivity analyses varying definitions of resistance or *Mtb* infection

**Supplemental Figure S6a:** Odds Ratios and confidence intervals for chromosome 13 locus rs1295104126 based on differing definitions of resistance in Sensitivity Analyses 1 & 2

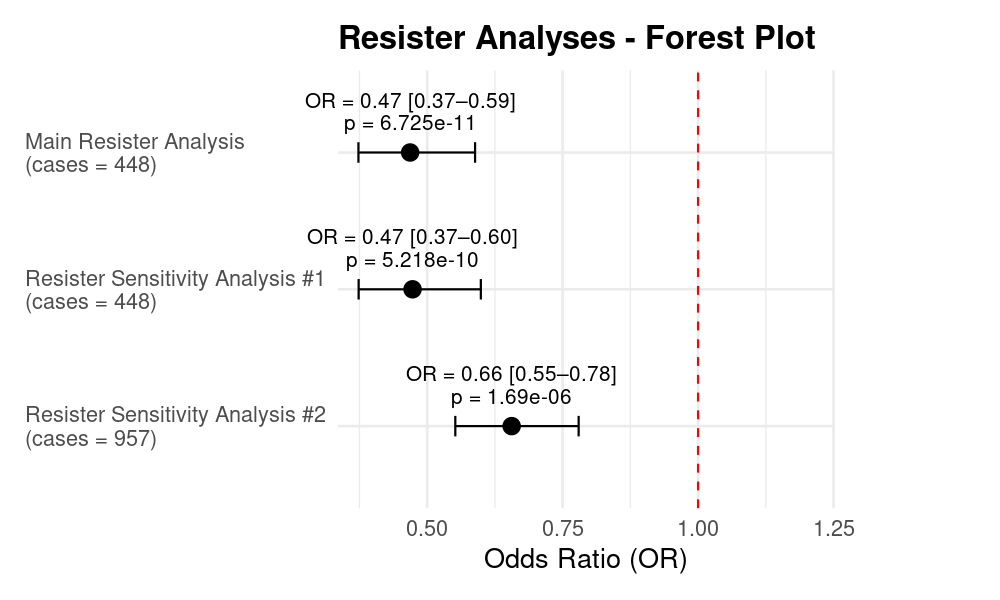

**Supplemental Figure S6b:** Odds Ratios and confidence intervals for chromosome 6 locus rs28752534 based on differing definitions of *Mtb* infection in Sensitivity Analysis 3

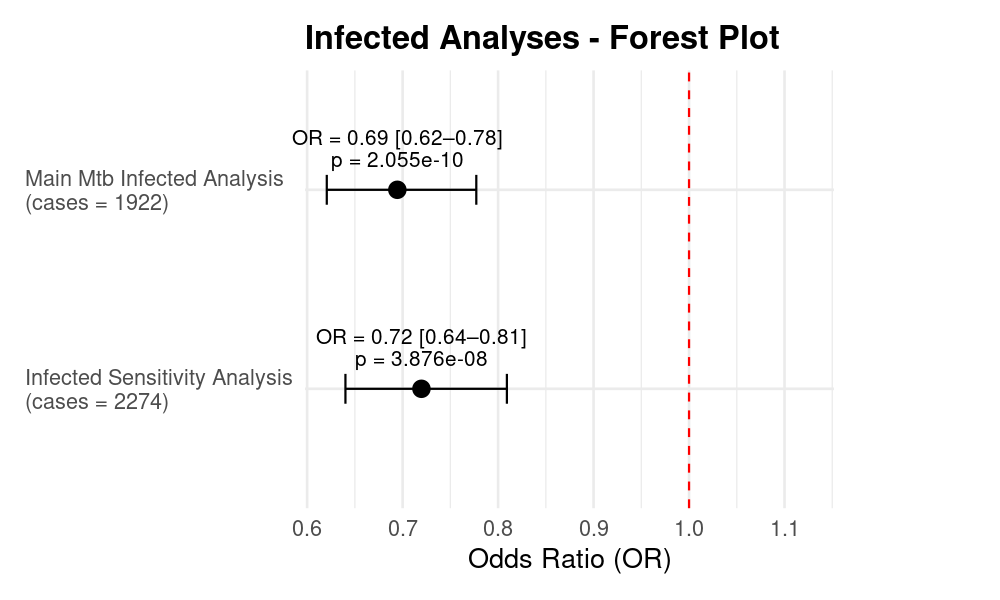
